# Predicting Hospital Admissions Using Pretrained EHR Embeddings: External Evaluation and Insights on Local Vocabulary Adaptation

**DOI:** 10.1101/2025.09.08.25335368

**Authors:** Bernardo Neves, Jorge Cerejo, Simão Gonçalves, Inês Mota, José M. Moreira, Nuno A. Silva, Francisca Leite, Michael Wornow, Mário J. Silva

## Abstract

**Purpose:** Unplanned hospital admissions impose substantial strain on healthcare systems, yet predictive models for these events remain underexplored in practice. This study evaluates whether publicly available pretrained transformer-based embeddings, developed on an external health system, can improve prediction of hospital admissions—including unplanned cases—when applied to a different institution with sparser data and a distinct medical vocabulary.

**Methods:** We performed a retrospective cohort study using structured EHR data from 200,000 adult patients (2007–2023) at a Portuguese hospital, standardized to the OMOP Common Data Model. Four 30-day outcomes were predicted: emergency department visits, hospital admissions, unplanned admissions, and readmissions. Three modeling approaches were compared: (1) clinically curated handcrafted features, (2) frequency-based representations of all recorded OMOP concepts, and (3) pretrained CLMBR-T embeddings generated from longitudinal OMOP data of 2.57 million patients in a U.S. hospital system. Performance was assessed on held-out patients using AUROC, AUPRC, and calibration metrics, with additional analysis of the impact of vocabulary overlap between pretraining and local datasets.

**Results:** Pretrained embeddings achieved the highest discrimination for all outcomes, particularly for unplanned admissions (AUROC 0.877 vs. 0.770 for counts). Gains were greatest for rarer outcomes and patients with richer clinical histories. Despite only 58% overlap with local vocabulary and substantially fewer events per patient than in pretraining, embeddings transferred effectively, indicating generalizable temporal patterns. Calibration was poorer than simpler models, necessitating post-hoc recalibration before deployment.

**Conclusion:** Pretrained OMOP-based EHR embeddings can substantially improve prediction of hospital and unplanned admissions in data- and resource-limited settings, even with partial vocabulary overlap. These findings support their use for rapid, cost-effective deployment of clinically meaningful predictive models, provided local recalibration and workflow integration are addressed.

**Highlights:**

- Superior cross-institution performance – Pretrained EHR embeddings from Stanford Medicine achieved AUROC 0.877 for unplanned admissions, 0.814 for hospital admissions, 0.782 for ED visits, and 0.923 for readmissions in a Portuguese hospital, outperforming count-based (0.770, 0.767, 0.744, 0.922) and handcrafted feature models across all tasks.
- Largest gains for rare, unpredictable events – For unplanned admissions (0.4% prevalence), embeddings nearly tripled AUPRC compared to counts (0.037 vs. 0.011) and improved AUROC by 0.107, with performance continuing to scale with more training data, unlike baselines.
- Effective under substantial domain shift – Strong transferability observed despite only 58% vocabulary overlap and markedly different patient populations, coding distributions, and event density (707 vs. 71 events per patient).
- Benefit increases with richer patient histories – Performance advantage of embeddings widened in patients with higher code volumes; AUROC for hospital admissions rose from 0.694 in the lowest quartile (Q1) to 0.870 in the highest (Q4), outperforming counts by up to 0.105 in Q4.
- Actionable guidance for adoption – Hospitals with sparse data can achieve rapid, cost-effective deployment of predictive models using external embeddings, especially for rare outcomes, if paired with local fine-tuning and post-hoc recalibration to ensure accurate risk estimation before clinical use.

## 1. Introduction

Unplanned hospital visits, including emergency department presentations and non-elective hospitalizations, impose substantial burdens on healthcare systems worldwide (Buja et al., 2020). Despite routine clinical risk assessment, healthcare providers typically rely on implicit judgment rather than validated, systematic tools for identifying high-risk patients during clinical encounters (Klunder et al., 2024). Traditional risk stratification instruments demonstrate limited predictive performance, with discriminative ability ranging from 0.61 to 0.78 AUC (Klunder et al., 2022), highlighting the need for more effective approaches.

Electronic Health Records (EHRs) contain rich longitudinal patient data with significant potential for dynamic risk prediction (Knevel and Liao, 2023). Recent advances in deep learning have demonstrated superior performance compared to traditional approaches, with transformer architectures emerging as the state-of-the-art for patient representation due to their ability to capture long-range dependencies in patient timelines (Si et al., 2021). Self-supervised learning has proven particularly valuable given the scarcity of labeled clinical data (Krishnan et al., 2022).

Foundational models—large-scale architectures trained on vast datasets through self-supervision—have emerged as powerful tools for EHR analysis that can adapt to multiple downstream tasks with minimal fine-tuning (Wornow et al., 2023b). These models offer improved performance, enhanced sample efficiency, and the ability to transfer learned representations across diverse healthcare settings, enabling external validation and improving prediction in data-constrained environments (Rasmy et al., 2021; Renc et al., 2024).

Transfer learning from foundational models addresses the significant costs and privacy barriers associated with developing models from scratch at each institution, with studies demonstrating that pretrained models can maintain 90% of optimal performance using only 30% of site-specific training data (Guo et al., 2024).

While the benefits of foundational models for clinical prediction have been established, their effectiveness for predicting unplanned visits has yet to be systematically evaluated, particularly regarding their ability to transfer knowledge from large datasets rich in vocabulary to smaller institutional settings. To address this gap, we investigate CLMBR-T-base, an autoregressive foundational model containing 141 million parameters pretrained on 2.57 million standardized patient records from the Observational Medical Outcomes Partnership (OMOP) Common Data Model (Wornow et al., 2023a). CLMBR-T-base was selected because it is currently the only publicly available foundational model with open weights specifically pretrained on longitudinal EHR data, making it particularly suitable for systematic evaluation across diverse healthcare institutions. Our study aims to determine whether the learned representations from this model can successfully adapt to the distinct challenge of predicting unplanned visits in healthcare environments with significantly different data volumes and vocabulary coverage. This scenario reflects real-world constraints faced by many healthcare institutions, where foundational models hold substantial promise for enhancing predictive capabilities.

## 2. Methods

This retrospective cohort study aimed to develop and compare prediction models for different types of hospital visits using historic EHR data from a single hospital. We compared three distinct visit-level feature strategies: 1) few simple handcrafted engineered features; 2) counts of all documented codes and 3) fixed vector representation derived from a pre-trained transformer model, (CLMBR-T-base) (Wornow et al., 2023a). All these approaches use all existing data about the patient up to the prediction time, only differing in the number of features and complexity of feature representation. The study diagram is depicted in Figure 1.

**Figure 1:**
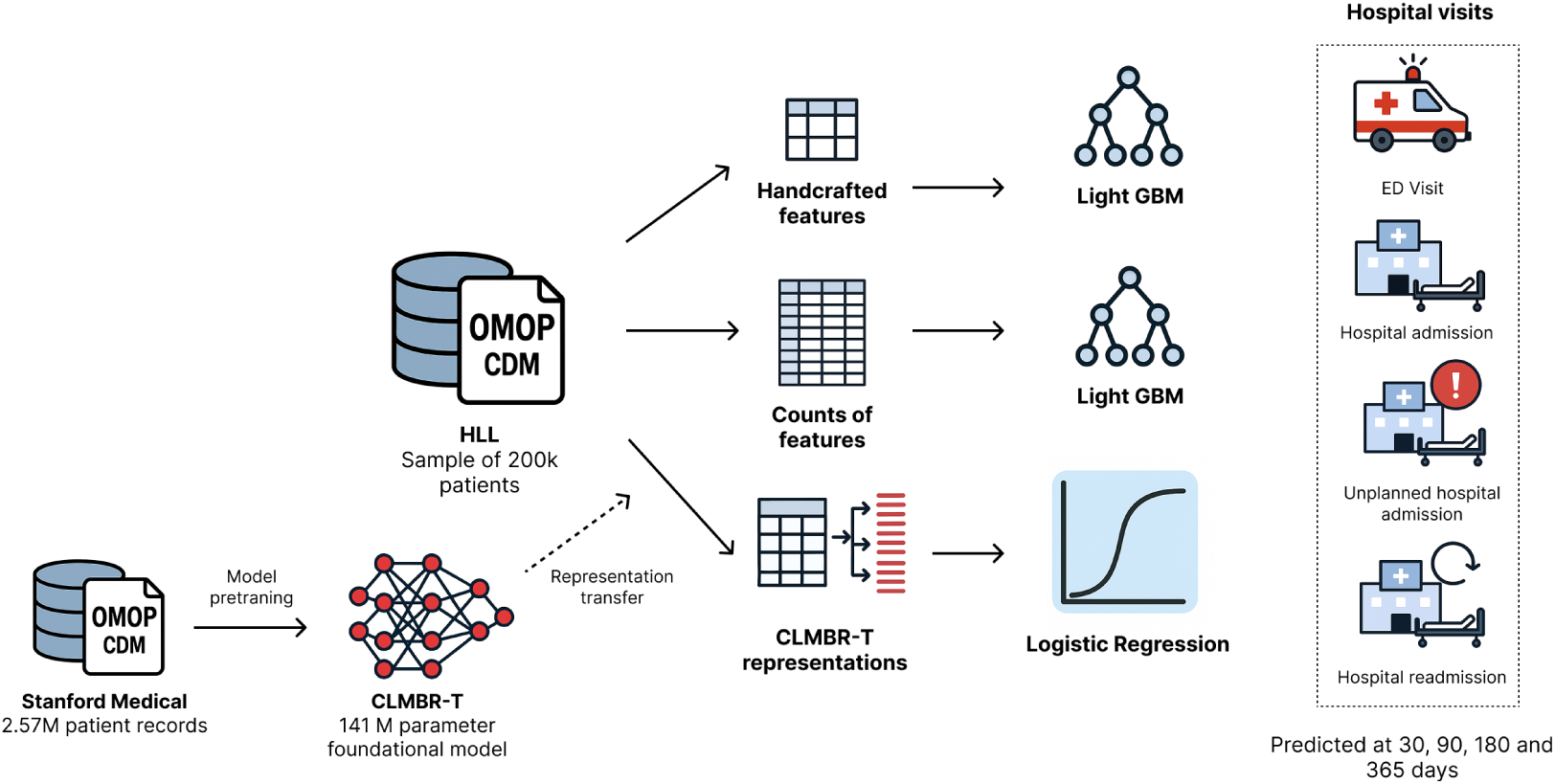
Graphical abstract of the study. Transfer learning workflow from large-scale pretrained CLMBR-T model (2.57M patients) to an external institutional dataset (200K patients) with 58% vocabulary overlap, led to superior performance across hospital visit prediction tasks compared to traditional feature-based approaches.

### 2.1. Data Source and Preprocessing

The data source for this study is a single hospital EHR database. Data were initially extracted from various clinical systems and transformed into the OMOP CDM version 5.4 (Reich et al., 2024). Specifically, this study utilized structured data from the person, visit occurrence, drug exposure, measurement, procedure occurrence, and condition occurrence tables. The dataset encompasses all hospital data from 2007 to 2023. A preliminary data quality assessment was conducted, followed by a data cleaning step where patients with missing identifiers, implausible demographics or temporal inconsistencies in their records were excluded.

### 2.2. Cohort Selection

Inclusion criteria required patients to be aged 18 years or older at their first recorded event and to have at least two documented hospital episodes of any type (outpatient, inpatient, or emergency department visits) within the study period. Each patient visit represented a potential prediction opportunity for forecasting future visits, with the exception of each patient’s final visit due to censoring.

We found 650,586 adult patients with at least two visits, and from this population we randomly sampled 200,000 patients for this study. This approach generated 2,714,826 prediction points across the entire dataset. We performed patient-level data splitting to ensure no patient appeared in multiple partitions, allocating 100,000 patients for model training, 50,000 for validation, and 50,000 for testing. Patient characteristics and split distributions summarized in Tables 1 and B.6, respectively.

### 2.3. Outcomes

At each prediction point, we predicted the occurrence of the following outcomes in the subsequent 30 days: 1) Emergency Department (ED) visit; 2) Any hospital admission; 3) Unplanned hospital admission: defined as an hospital admission that was preceded by an ED visit in the previous 48 hours; 4) Hospital readmission: defined as a second hospital admission after a first index admission.

### 2.4. Patient representations

At each prediction point, three distinct representations of patients patients were created and compared.

1. **Handcrafted features**: Based on prior literature (Askar et al., 2024), we developed handcrafted features relevant for predicting unplanned hospital admissions. We transformed OMOP CDM data into a visit-level matrix of dimensions *n*^′^ *× d*, where *n*^′^ represents all patient visits excluding each patient’s final visit, and *d* is the number of features. Features were engineered using information available up to each visit and included: 1) patient demographics (gender and age at visit); 2) healthcare utilization metrics (cumulative counts of outpatient visits, ED visits, total and distinct prescribed drugs); and 3) multimorbidity encoded as the Charlson Comorbidity Index (Lee et al., 2021), calculated using locally developed phenotyping rules that mapped clinical concepts to diseases (Neves et al., 2025).
2. **Counts of previous concepts**: We implemented count-based featurization as a competitive baseline following established best practices (Rajkomar et al., 2018). Our OMOP CDM data was first converted to the Medical Event Data Standard (MEDS) format to enable standardized processing. We then used the FEMR (Framework for Electronic Medical Records) ^1^ package to generate count-based features, which transforms each patient into a feature vector where each element represents the frequency of a specific clinical concept occurring in the patient’s timeline prior to the prediction time point. The FEMR CountFeaturizer was configured to include string value combinations (incorporating laboratory results and other coded values) and age normalization features, creating a comprehensive representation of each patient’s historical clinical encounters.
3. **CLMBR-T representations**: We utilized CLMBR-T-base as our foundational model to generate patient representations from our data. This publicly available 141-million-parameter autoregressive Transformer model was pre-trained on sequences of clinical codes (e.g., diagnosis, procedure, medication, lab result code) to predict the next medical code, enabling it to learn rich representations of patient trajectories. The model was originally pre-trained on the large-scale, de-identified Stanford Medicine Research Data Repository (STARR) dataset (Datta et al., 2020), which is also in the OMOP CDM format and contains structured EHR data. Detailed model training specifications can be found in the original model paper (Wornow et al., 2023a). To create these representations we first converted data in all OMOP tables into the Medical Event Data Standard (MEDS) format, a data schema for storing streams of medical events that is increasingly being employed for EHR predictive modeling (Arnrich et al., 2024). Given that converting the whole dataset would require expensive computational and storage costs we sampled 100,000 patients from each of the train and test splits using stratified sampling to maintain the original outcome distribution. We used the FEMR tokenizer and FEMR labeler from FEMR package to convert patient data into token sequences and to create the desired labels for each prediction. These were given as input to the CLMBR-T-base model that generated fixed 768-dimensional vector representations (embeddings) for each patient at the specified prediction timepoints. These embeddings encapsulate the patient’s clinical history as learned by the transformer through its pre-training on longitudinal medical data, incorporating both the temporal sequence of medical events and continuous values such as laboratory results Wornow et al. (2023a). A comparison of the codes used for creating representations in pretrained model and in our dataset are provided in Tables 5 and B.10.

### 2.5. Modeling

For the handcrafted feature and count-based representations, we implemented a machine learning pipeline using LightGBM, a gradient boosting framework selected for its demonstrated effectiveness on tabular data, built-in class imbalance handling capabilities, and use as the primary baseline in the original CLMBR-T paper, ensuring direct methodological comparability (Wornow et al., 2023a). Our dataset exhibited considerable class imbalance across outcomes at 30 days (ED visits: 5.87%, hospital admissions: 8.32%, unplanned admissions: 0.47%, readmissions: 24.18%). Therefore we employed LightGBM with automatic class imbalance handling and hyper-parameters optimized through grid search over learning rates of [0.02, 0.1, 0.2-0.5], maximum depths of [3, 5-6, 7], and number of leaves [5-10, 15-25, 31-100], with early stopping (50 rounds) and other parameters including feature fraction (0.8), bagging fraction (0.8), and bagging frequency (5) to address the severe class imbalance typical in healthcare prediction tasks. For count-based representations, we conducted experiments across multiple training sample sizes (5k, 10k, 25k, 50k, and all 100k available patients) using a patient-centric sampling approach where all visits for each selected patient were included, maintaining longitudinal trajectory information while preserving original outcome prevalence through stratified sampling. To mitigate the influence of high-utilization patients, we implemented a 32-visit cap per patient, randomly sampling visits when this threshold was exceeded to preserve average patient information. For handcrafted feature experiments, we utilized all available pre-assigned training patients without sub-sampling to provide the strongest possible baseline for representation comparison. Data preprocessing included LightGBM’s native missing value handling supplemented by domain-informed imputation strategies, with consistent application of the visit cap across both representation types to ensure fair comparison.

For the pretrained feature representation strategy, we followed the original CLMBR-T methodology by fine-tuning CLMBR-T-base models using few-shot learning for each prediction task Wornow et al. (2023a). The finetuning process employed outcome-stratified patient sampling with varying training sizes (5k, 10k, 25k, 50k, and all available training patients), mirroring the count-based approach to enable direct comparison. Patient-centric sampling was implemented to preserve longitudinal trajectory information, where all visits for each selected patient were included up to the 32-visit cap, maintaining the temporal sequence crucial for clinical prediction. After fine-tuning, the resulting CLMBR-T representations (768-dimensional vectors) were frozen and used as fixed feature inputs to train downstream logistic regression classifiers with L2 regularization (C=1.0) and balanced class weights to address the natural class imbalance in healthcare prediction tasks. The same data split was employed, with final evaluation on 50k held-out test patients, ensuring direct comparability with handcrafted and count-based results (Figure 1).

### 2.6. Evaluation Framework

All models were evaluated using a comprehensive set of metrics appropriate for imbalanced healthcare prediction tasks. Discrimination performance was assessed via AUROC (Area Under Receiver Operating Characteristic curve) and AUPRC (Area Under Precision-Recall Curve). Classification performance was evaluated through F1 score, precision, recall, and specificity at an optimal threshold determined by maximizing F1 score on the validation set. The optimal threshold was identified by calculating F1 scores across all possible thresholds derived from the precision-recall curve and selecting the threshold that achieved the highest F1 score, ensuring balanced performance across both positive and negative classes—a critical consideration given the severe class imbalance typical in clinical prediction scenarios (Hicks et al., 2022).

Model calibration was quantified using Expected Calibration Error (ECE, computed as the average absolute difference between predicted probabilities and actual frequencies across 10 probability bins), Maximum Calibration Error (MCE, the maximum absolute difference across all bins), and Brier score (mean squared difference between predicted probabilities and actual binary outcomes). All threshold optimization was performed exclusively on the validation set (50k patients), with final performance assessment conducted on the completely held-out test set (50k patients) to prevent overfitting and ensure unbiased evaluation.

To quantify uncertainty while respecting the study’s hierarchical design, we computed 95% confidence intervals for AUROC and AUPRC using a patient-level cluster bootstrap with 1,000 replicates. In each iteration we resampled whole patients with replacement, retained all of their encounters, recalculated the metric, and formed the interval from the 2.5th and 97.5th percentiles of the empirical distribution. By resampling patients rather than visits, this cluster bootstrap approach preserves within-patient correlation, avoids anticonservative bias, remains assumption-lean, and achieves near-nominal coverage across diverse hierarchical data settings, making it well suited for our multi-visit clinical dataset (Ren et al., 2010).

### 2.7. Software libraries

All data preprocessing, feature engineering, modeling, and evaluation tasks were conducted using *python*, leveraging specialized packages: pandas, polars, and scikit-learn. For using CLMBR-T-base we converted OMOP data to MEDS format using meds-etl package ^2^. The FEMR package was used to handle data, more specifically create labels and tokenize, before data was processed by the model. Model weights were downloaded from the Huggingface repository ^3^.

## 3. Results

### 3.1. Dataset Description and Outcome Prevalence

The study cohort comprised 200,000 adult patients with a mean age of 48.5 years (SD 17.7), of whom 57.8% were female. The population demonstrated moderate healthcare utilization patterns, with nearly half (45.8%) having at least one emergency department visit and approximately one-quarter (23.8%) experiencing hospital admissions during the study period. Outpatient care was nearly universal, with 95.8% of patients having at least one outpatient visit (median 5 visits per patient). Acute care events requiring readmission or unplanned admission were relatively uncommon, affecting only 2.1% and 2.8% of patients respectively. The burden of chronic conditions was moderate, with 39.3 of patients having at least one documented chronic condition and 17.5% exhibiting multimorbidity (two or more conditions).

Four distinct 30-day outcome prediction tasks were evaluated: ED visits, hospital admissions, readmissions at 30 days, and unplanned admissions. Each patient generated multiple prediction points at the end of each clinical encounter, except for the last observable one. For readmissions, predictions were made only at hospital discharge events. This approach yielded 8,361,646 total prediction opportunities across all prediction tasks. The prevalence of positive outcomes varied substantially across tasks, ranging from 0.4% for unplanned admissions to 24.8% for readmissions at 30 days, reflecting the different risk profiles and clinical contexts of each prediction scenario (Table 1).

**Table 1:** Population Characteristics and Prediction Task Overview.

| Population Characteristics |  |  |
| --- | --- | --- |
| Total Patients | 200,000 |  |
| Mean Age (SD), years | 48.5 (17.7) |  |
| Female, n (%) | 115,693 (57.8) |  |
| Total Visits | 2,896,185 |  |
| Visits per Patient | 14.5 |  |
| Healthcare Utilization |  |  |
| ED Visits |  |  |
| Patients with $\geq 1$, n (%) | 91,675 (45.8) | |
| Total visits | 275,616 |  |
| Per patient (median [IQR]) | 0 [0-2] |  |
| Hospital Admissions |  |  |
| Patients with $\geq 1$, n (%) | 47,640 (23.8) | |
| Total visits | 90,999 |  |
| Per patient (median [IQR]) | 0 [0-0] |  |
| Readmissions at 30 days |  |  |
| Patients with $\geq 1$, n (%) | 4,259 (2.1) | |
| Total visits | 20,893 |  |
| Per patient (median [IQR]) | 0 [0-0] |  |
| Unplanned Admissions |  |  |
| Patients with $\geq 1$, n (%) | 5,679 (2.8) | |
| Total visits | 6,593 |  |
| Per patient (median [IQR]) | 0 [0-0] |  |
| Outpatient Visits |  |  |
| Patients with $\geq 1$, n (%) | 191,632 (95.8) | |
| Total visits | 2,529,570 |  |
| Per patient (median [IQR]) | 5 [2-13] |  |
| Chronic Conditions |  |  |
| Patient with $\geq 1$ Chronic Condition, n (%) | 78,529 (39.3) | |
| Multimorbidity, n (%) | 34,980 (17.5) |  |
| Prediction Tasks |  |  |
|  | Prediction Points | Positive Cases (%) |
| ED Visits | 2,696,185 | 156,024 (5.8) |
| Hospital Admissions | 2,709,596 | 227,363 (8.4) |
| Readmissions | 252,956 | 62,597 (24.8) |
| Unplanned Admissions | 2,702,909 | 10,067 (0.4) |

### 3.2. Performance Comparison Between Model Approaches

Performance varied substantially across the three modeling approaches, with CLMBR-T consistently outperforming count-based features, which in turn exceeded handcrafted features across all prediction tasks (Table 2 and Figure 2).

**Figure 2:**
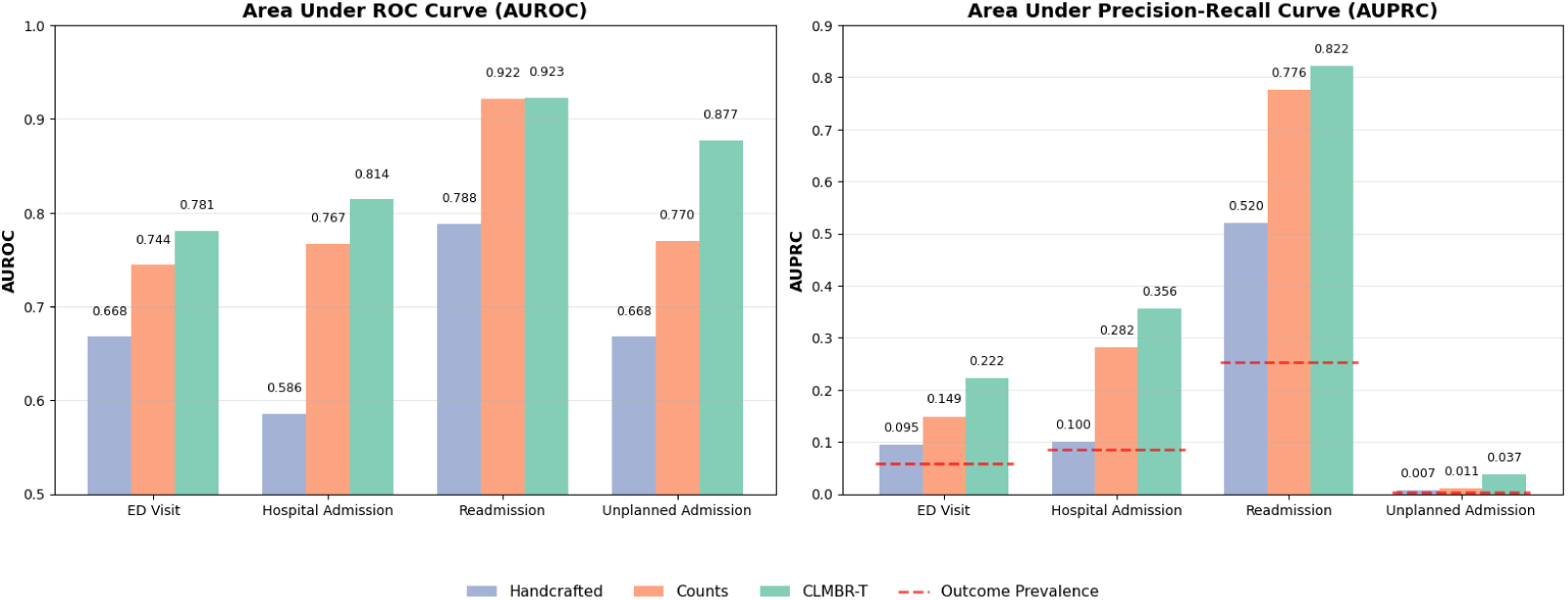
Performance comparison of three modeling approaches across four clinical prediction tasks. The left panel shows Area Under the Receiver Operating Characteristic Curve (AUROC) values, while the right panel displays Area Under the Precision-Recall Curve (AUPRC) values for each outcome. Three modeling strategies are compared: hand-crafted features (light blue), counts-based features (orange), and CLMBR-T deep learning embeddings (green). Red dashed lines in the AUPRC panel indicate outcome prevalence baselines.

**Table 2:** Performance of Handcrafted, Counts, and CLMBR-T Models on Four Outcomes (Test Set).

| Outcome (Prevalence) | Metric | Handcrafted | Counts | CLMBR-T |
| --- | --- | --- | --- | --- |
| ED Visit (5.8%) | AUROC | 0.668 | 0.744 (0.738–0.749) | 0.782 (0.777–0.786) |
|  | AUPRC | 0.095 | 0.150 (0.144–0.155) | 0.222 (0.214–0.229) |
|  | Recall | 0.360 | 0.408 | 0.347 |
|  | Specificity | 0.838 | 0.878 | 0.935 |
|  | Precision | 0.110 | 0.152 | 0.232 |
|  | NPV | 0.960 | 0.965 | 0.962 |
|  | F1 Score | 0.168 | 0.222 | 0.278 |
| Hospital Admission (8.5%) | AUROC | 0.586 | 0.767 (0.761–0.773) | 0.814 (0.809–0.819) |
|  | AUPRC | 0.100 | 0.282 (0.268–0.296) | 0.356 (0.341–0.372) |
|  | Recall | 0.230 | 0.366 | 0.402 |
|  | Specificity | 0.877 | 0.943 | 0.954 |
|  | Precision | 0.126 | 0.329 | 0.382 |
|  | NPV | 0.937 | 0.952 | 0.958 |
|  | F1 Score | 0.163 | 0.347 | 0.392 |
| Readmission (25.3%) | AUROC | 0.788 | 0.922 (0.915–0.928) | 0.923 (0.915–0.930) |
|  | AUPRC | 0.520 | 0.777 (0.750–0.800) | 0.822 (0.799–0.841) |
|  | Recall | 0.667 | 0.778 | 0.709 |
|  | Specificity | 0.769 | 0.906 | 0.951 |
|  | Precision | 0.451 | 0.704 | 0.808 |
|  | NPV | 0.890 | 0.935 | 0.919 |
|  | F1 Score | 0.538 | 0.739 | 0.755 |
| Unplanned Admission (0.4%) | AUROC | 0.668 | 0.770 (0.744–0.795) | 0.877 (0.865–0.890) |
|  | AUPRC | 0.007 | 0.011 (0.010–0.013) | 0.037 (0.032–0.044) |
|  | Recall | 0.066 | 0.396 | 0.152 |
|  | Specificity | 0.979 | 0.909 | 0.991 |
|  | Precision | 0.011 | 0.014 | 0.061 |
|  | NPV | 0.997 | 0.998 | 0.997 |
|  | F1 Score | 0.019 | 0.028 | 0.087 |

**Notes:** AUROC = Area Under the Receiver Operating Characteristic Curve; AUPRC = Area Under the Precision–Recall Curve; PPV = Positive Predictive Value (Precision); NPV = Negative Predictive Value. Values in parentheses are 95% confidence intervals. Optimal thresholds determined by maximizing F1 score on the validation set.

CLMBR-T demonstrated superior discriminative performance, achieving the highest AUROC values for all outcomes: 0.782 for ED visits, 0.814 for hospital admissions, 0.923 for readmissions, and 0.877 for unplanned admissions. Count-based features showed intermediate performance with AU-ROCs of 0.744, 0.767, 0.922, and 0.770, respectively, while handcrafted features achieved the lowest discrimination with AUROCs ranging from 0.586 to 0.788.

The performance advantages of CLMBR-T were particularly pronounced for precision-recall metrics and rare events. For unplanned admissions (0.4% prevalence), CLMBR-T achieved an AUPRC of 0.037 compared to 0.011 for counts and 0.007 for handcrafted features. Readmissions showed the smallest performance gaps, with count-based and CLMBR-T models achieving nearly identical AUROC values (0.922 vs 0.923).

Model calibration revealed a trade-off between discriminative performance and probability calibration. Handcrafted features demonstrated excellent calibration (ECE *geq* 0.075), while CLMBR-T models showed substantial miscalibration (ECE: 0.193-0.486), and count-based models exhibited intermediate calibration performance (ECE: 0.002-0.356). Detailed metrics in Table B.7.

### 3.3. Train Sample Size Efficiency Analysis

We systematically evaluated the effect of training sample size on model performance by training both count-based and CLMBR-T models with varying numbers of patients (5k, 10k, 25k, 50k, and 100k). The results demonstrate distinct scaling patterns between the two approaches across all prediction tasks (Table 3 and Figure 3).

**Figure 3:**
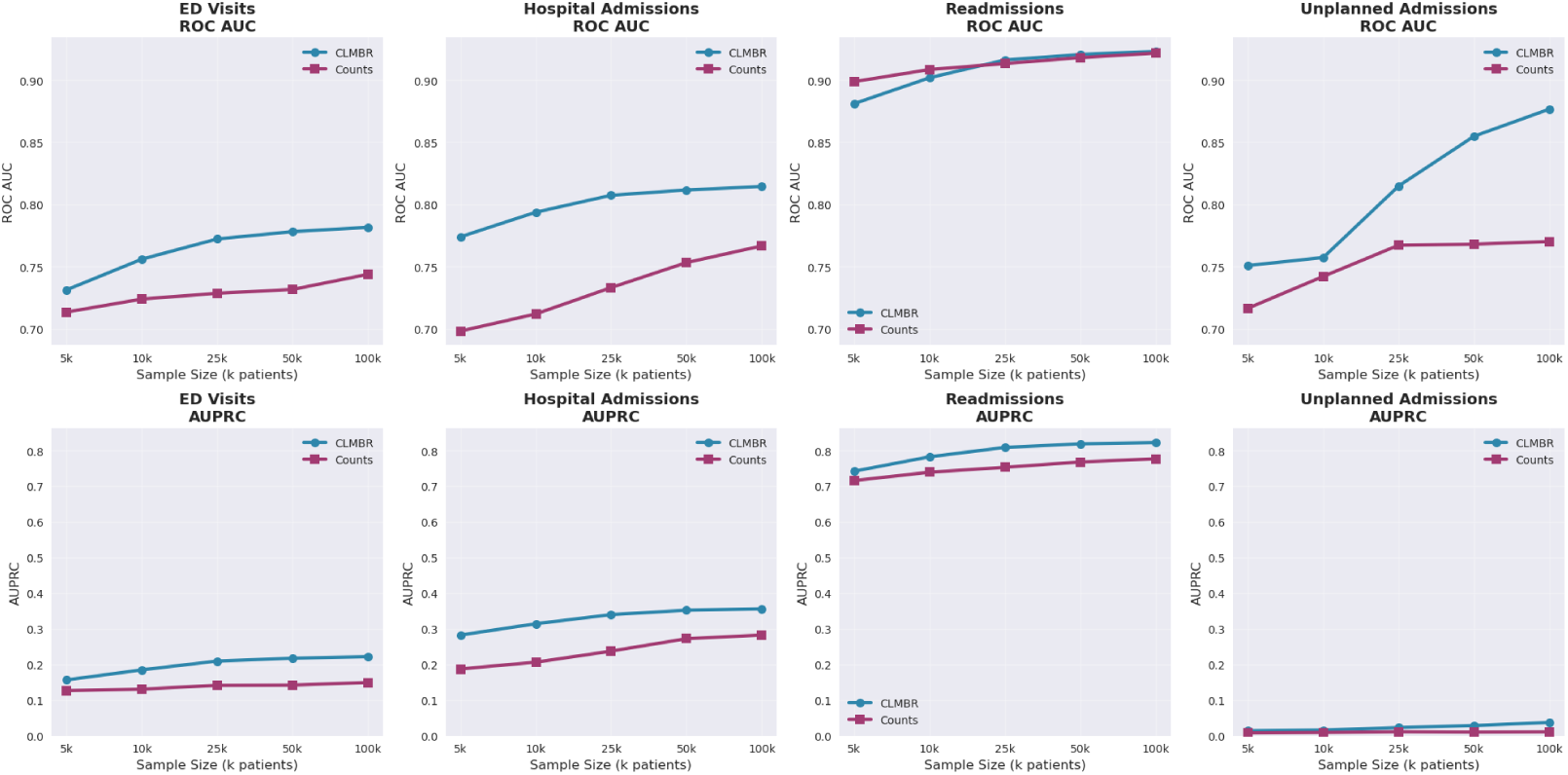
Performance comparison between CLMBR-T and counts-based models across different patient sample sizes. Top row shows ROC AUC performance, bottom row shows AUPRC performance. CLMBR-T consistently outperforms counts-based models across most tasks, with the largest improvements observed for unplanned admissions.

**Table 3:** Impact of Training Sample Size on Model Performance Across Prediction Tasks.

| Outcome | Method | Training Sample Size |  |  |  |  |
| --- | --- | --- | --- | --- | --- | --- |
|  |  | 5k | 10k | 25k | 50k | All (100k) |
| ED Visits | Count-based | 0.713 (0.127) | 0.728 (0.128) | 0.732 (0.147) | 0.738 (0.148) | 0.744 (0.149) |
|  | CLMBR-T | 0.731 (0.154) | 0.756 (0.185) | 0.772 (0.210) | 0.778 (0.218) | 0.782 (0.222) |
| Hospital Admissions | Count-based | 0.699 (0.184) | 0.712 (0.207) | 0.733 (0.235) | 0.752 (0.267) | 0.767 (0.282) |
|  | CLMBR-T | 0.774 (0.283) | 0.794 (0.314) | 0.807 (0.341) | 0.812 (0.352) | 0.814 (0.356) |
| Readmissions | Count-based | 0.900 (0.720) | 0.908 (0.743) | 0.913 (0.752) | 0.918 (0.770) | 0.922 (0.776) |
|  | CLMBR-T | 0.881 (0.742) | 0.902 (0.783) | 0.916 (0.810) | 0.921 (0.817) | 0.923 (0.822) |
| Unplanned Admissions | Count-based | 0.718 (0.009) | 0.754 (0.010) | 0.767 (0.011) | 0.770 (0.011) | 0.770 (0.011) |
|  | CLMBR-T | 0.751 (0.014) | 0.757 (0.016) | 0.815 (0.024) | 0.855 (0.029) | 0.877 (0.037) |

Results show AUROC (AUPRC) for each training sample size. CLMBR-T = CLMBR-T-base transformer model; Count-based = frequency-based feature representation. Values represent mean performance across multiple runs where available.

Count-based features showed consistent but modest improvements with increased training data across most outcomes. For ED visits, AUROC improved from 0.713 (5k patients) to 0.744 (100k patients), representing a 4.3% relative improvement. Hospital admissions demonstrated the largest gains, with AUROC increasing from 0.699 to 0.767 (9.7% improvement). Readmissions showed steady but smaller improvements (0.900 to 0.922, 2.4% gain), while unplanned admissions reached a performance plateau at 25k patients, with minimal improvement beyond this point (0.767 to 0.770).

The transformer-based approach exhibited more pronounced scaling benefits, particularly for precision-recall performance. ED visits showed substantial improvement from 0.731 to 0.782 AUROC (7.0% gain), with AUPRC increasing from 0.154 to 0.222 (44% relative improvement). Hospital admissions demonstrated strong scaling from 0.774 to 0.814 AUROC, while AUPRC improved from 0.283 to 0.356 (26% gain). Notably, CLMBR-T showed the most dramatic scaling for unplanned admissions, with AUROC improving from 0.751 to 0.877 (16.8% gain) and AUPRC nearly tripling from 0.014 to 0.037.

Different prediction tasks exhibited varying sensitivity to training sample size. Readmissions, the most balanced outcome (22% prevalence), showed modest improvements for both approaches, with diminishing returns beyond 25k patients. In contrast, unplanned admissions (0.4% prevalence) demonstrated continued improvement with larger training sets, particularly for CLMBR-T models. This suggests that rare events benefit more substantially from increased training data, especially when using representation learning approaches. Count-based models generally reached performance plateaus earlier, with most improvements achieved by 25-50k patients. CLMBR-T models showed more linear scaling patterns, suggesting potential for further improvement with additional training data. The gap between approaches widened with larger training sets, indicating that transformer-based models can better exploit increased data availability.

### 3.4. Performance by Code Volume and Vocabulary Transfer Analysis

To evaluate the relationship between available historical information and model performance, we stratified test patients into quartiles based on the total number of clinical codes (including diagnoses, procedures, medications, and laboratory measurements) accumulated in their EHR up to each prediction timepoint (Q1: 2-5 codes, Q2: 6-27 codes, Q3: 28-79 codes, Q4: 80+ codes). The number of available codes varied dynamically across prediction instances for the same patient, reflecting the cumulative nature of clinical information available at different timepoints throughout their care trajectory (Table 4 and Figure 4).

**Figure 4:**
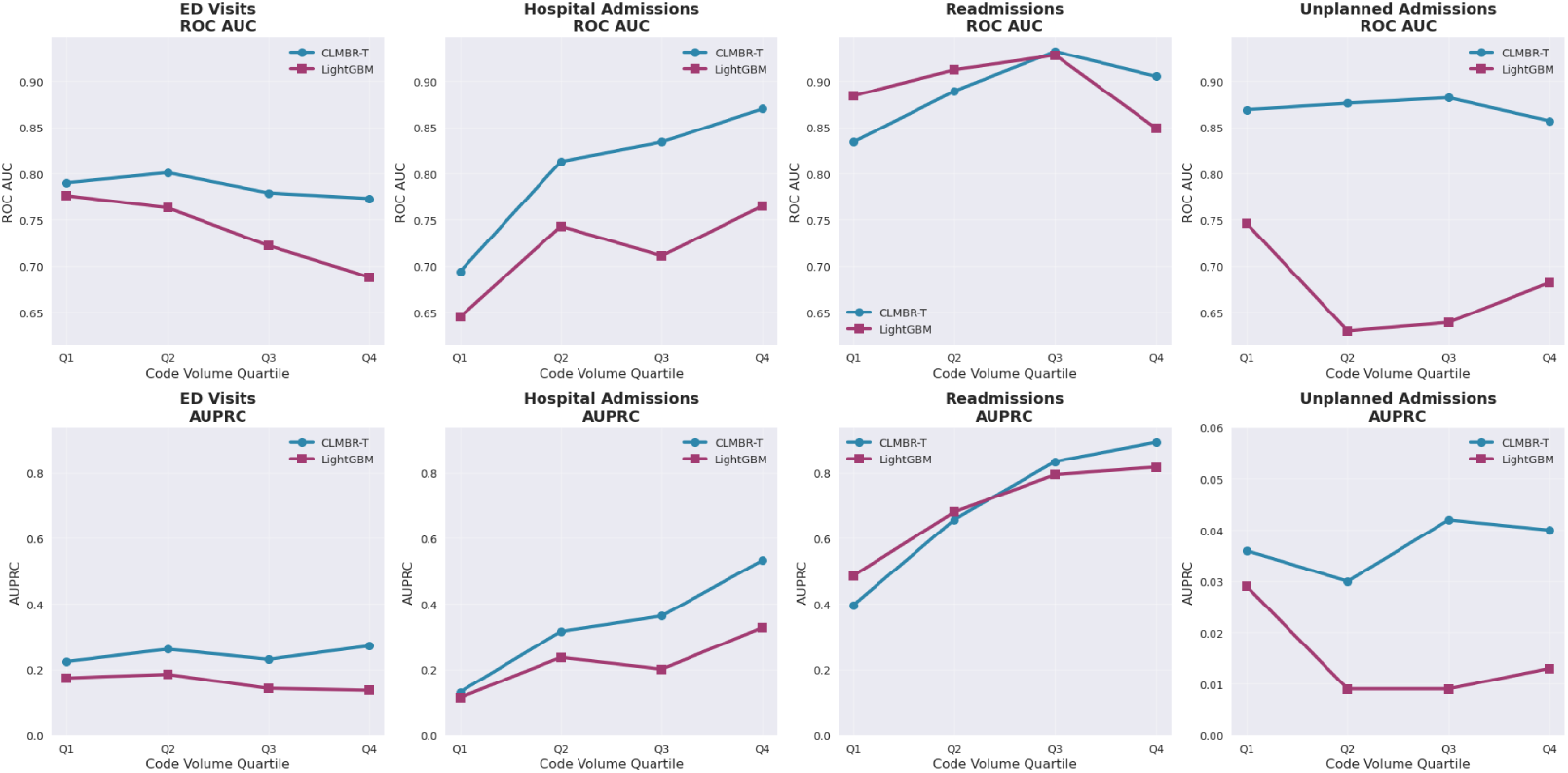
CLMBR-T and Count-based Models Performance Across Code Volume Quartiles in Test Set. Q1: 2-5, Q2: 6-27, Q3: 28-79, Q4: 80+ codes. Both approaches improve with higher code volumes, but CLMBR-T’s advantage widens substantially in higher quartiles. Performance gains are most pronounced for hospital admissions and readmissions, while ED visits and unplanned admissions show modest improvements. The expanding gap demonstrates CLMBR-T’s superior ability to leverage rich clinical histories.

**Table 4:**
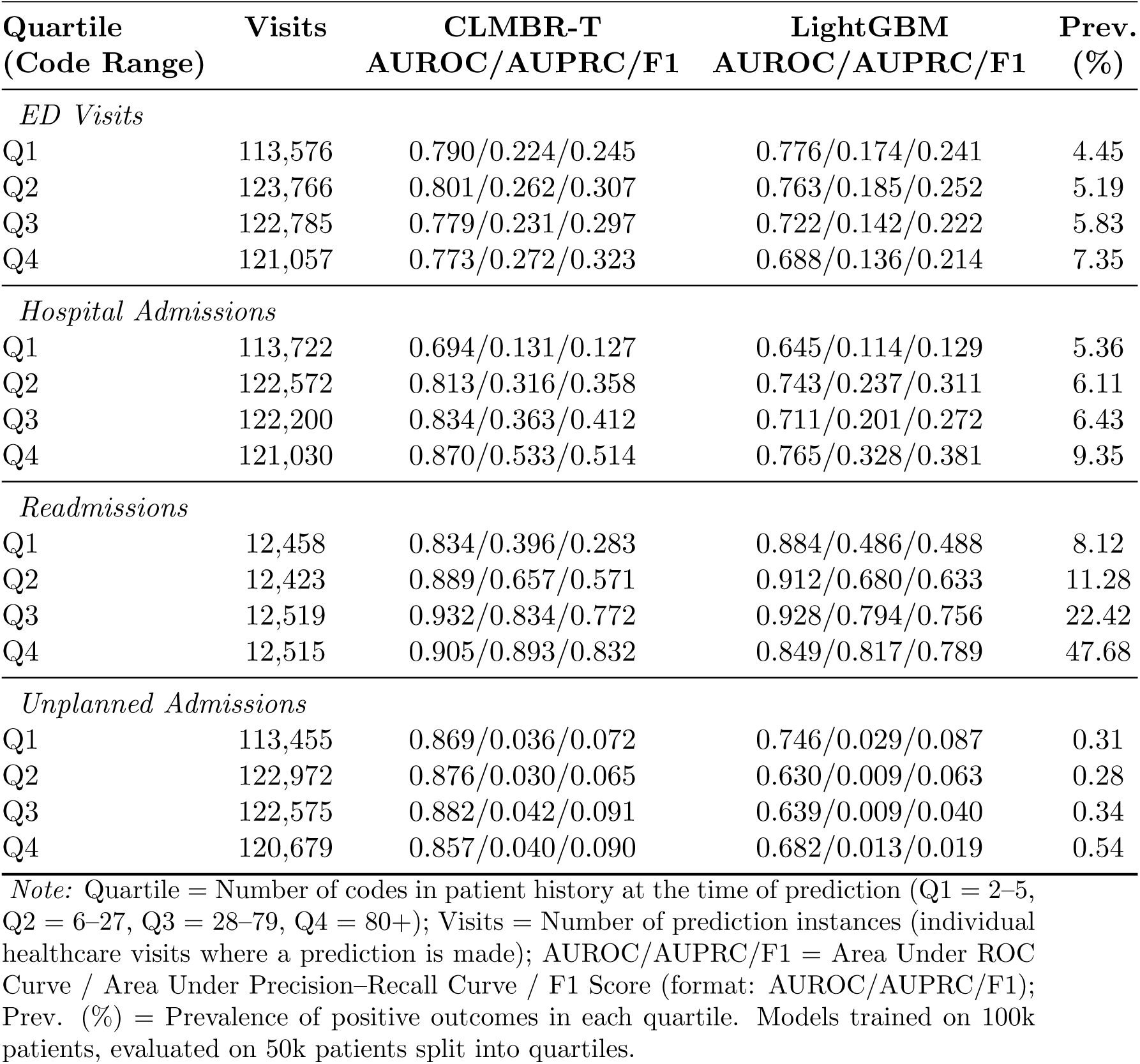
Model Performance Across Code Volume.

| Quartile<br>(Code Range) | Visits | CLMBR-T<br>AUROC/AUPRC/F1 | LightGBM<br>AUROC/AUPRC/F1 | Prev.<br>(%) |
| --- | --- | --- | --- | --- |
| *ED Visits* |  |  |  |  |
| Q1 | 113,576 | 0.790/0.224/0.245 | 0.776/0.174/0.241 | 4.45 |
| Q2 | 123,766 | 0.801/0.262/0.307 | 0.763/0.185/0.252 | 5.19 |
| Q3 | 122,785 | 0.779/0.231/0.297 | 0.722/0.142/0.222 | 5.83 |
| Q4 | 121,057 | 0.773/0.272/0.323 | 0.688/0.136/0.214 | 7.35 |
| *Hospital Admissions* |  |  |  |  |
| Q1 | 113,722 | 0.694/0.131/0.127 | 0.645/0.114/0.129 | 5.36 |
| Q2 | 122,572 | 0.813/0.316/0.358 | 0.743/0.237/0.311 | 6.11 |
| Q3 | 122,200 | 0.834/0.363/0.412 | 0.711/0.201/0.272 | 6.43 |
| Q4 | 121,030 | 0.870/0.533/0.514 | 0.765/0.328/0.381 | 9.35 |
| *Readmissions* |  |  |  |  |
| Q1 | 12,458 | 0.834/0.396/0.283 | 0.884/0.486/0.488 | 8.12 |
| Q2 | 12,423 | 0.889/0.657/0.571 | 0.912/0.680/0.633 | 11.28 |
| Q3 | 12,519 | 0.932/0.834/0.772 | 0.928/0.794/0.756 | 22.42 |
| Q4 | 12,515 | 0.905/0.893/0.832 | 0.849/0.817/0.789 | 47.68 |
| *Unplanned Admissions* |  |  |  |  |
| Q1 | 113,455 | 0.869/0.036/0.072 | 0.746/0.029/0.087 | 0.31 |
| Q2 | 122,972 | 0.876/0.030/0.065 | 0.630/0.009/0.063 | 0.28 |
| Q3 | 122,575 | 0.882/0.042/0.091 | 0.639/0.009/0.040 | 0.34 |
| Q4 | 120,679 | 0.857/0.040/0.090 | 0.682/0.013/0.019 | 0.54 |

*Note:* Quartile = Number of codes in patient history at the time of prediction (Q1 = 2–5, Q2 = 6–27, Q3 = 28–79, Q4 = 80+); Visits = Number of prediction instances (individual healthcare visits where a prediction is made); AUROC/AUPRC/F1 = Area Under ROC Curve / Area Under Precision–Recall Curve / F1 Score (format: AUROC/AUPRC/F1); Prev. (%) = Prevalence of positive outcomes in each quartile. Models trained on 100k patients, evaluated on 50k patients split into quartiles.

Outcome prevalence increased substantially with higher historical code volume for most prediction tasks. Readmissions demonstrated the strongest gradient, with prevalence increasing from 8.12% (Q1) to 47.68% (Q4). Hospital admissions showed a similar pattern, rising from 5.36% to 9.35% across quartiles. ED visits increased modestly from 4.45% to 7.35%, while unplanned admissions remained relatively stable (0.28-0.54%) across code volume levels.

CLMBR-T performance improved substantially with increasing historical code volume. For hospital admissions, AUROC increased from 0.694 (Q1) to 0.870 (Q4), while AUPRC improved from 0.131 to 0.533. Readmissions showed strong performance across all quartiles, with AUROC ranging from 0.834 to 0.932 and AUPRC from 0.396 to 0.893. ED visits maintained consistent discrimination (AUROC: 0.773-0.801) across code volume levels. Unplanned admissions achieved excellent discrimination across all quartiles (AUROC: 0.857-0.882). The performance advantage of CLMBR-T over count-based features increased with higher code volume. For hospital admissions, the AUROC difference widened from 0.049 (Q1) to 0.105 (Q4). Count-based features showed superior performance for readmissions in lower volume quartiles (Q1-Q2), with CLMBR-T gaining advantage only in higher volume patients (Q3-Q4).

Vocabulary analysis revealed substantial differences between the CLMBR-T pretraining dataset and our institutional data (Table 5). The pretrained model utilized 26,145 unique codes compared to 7,712 in our dataset, with 4,489 codes shared between datasets. This overlap represented 58.2% of our vocabulary but only 17.2% of the pretraining vocabulary. Shared codes consisted primarily of SNOMED concepts (90.5%), LOINC laboratory codes (6.9%), and RxNorm medication codes (2.5%). Despite this limited vocabulary overlap and substantially lower event density (71 vs 707 events per patient), CLMBR-T maintained strong transfer learning performance across all prediction tasks.

**Table 5:** Dataset Characteristics and Vocabulary Overview.

| Characteristic | CLMBR-T | Our Dataset |
| --- | --- | --- |
| *Dataset Characteristics* |  |  |
| Number of patients | 2,570,000 | 100,000 |
| Events per patient (mean) | 707 | 71 |
| Visits per patient (mean) | 28 | 18 |
| Min visits per patient | 0 | 1 |
| Max visits per patient | 3,701 | 893 |
| *Overall Vocabulary Statistics* |  |  |
| Total vocabulary size | 26,145 | 7,712 |
| Shared codes (overlap) | 4,489 (17.17%) | 4,489 (58.21%) |
| Dataset-specific codes | 21,656 (82.83%) | 3,223 (41.79%) |

*Note:* Shared codes refer to medical codes present in both datasets. Percentages indicate the proportion of each dataset’s vocabulary that is shared vs. dataset-specific. CLMBR-T data from (Wornow et al., 2023a); Our Dataset from training set. Detailed vocabulary breakdown available in Supplementary Table B.10.

## 4. Discussion

In this study, we evaluated the effectiveness of transfer learning using a publicly available foundational model (CLMBR-T-base) for predicting hospital visits in a new institutional setting characterized by significantly less comprehensive data compared to the model’s original training environment. Our analysis showed that despite notable differences in data density and vocabulary coverage, CLMBR-T-base with a logistic regression classifier consistently outperformed LightGBM models trained on either handcrafted features or counts of the entire data vocabulary across all evaluated outcomes. Our primary finding is that pretrained transformer models can generalize effectively across healthcare institutions, even when significant disparities exist in vocabulary size, overlap, and patient event density. Specifically, despite our institutional dataset containing approximately one-tenth of the event density per patient compared to the CLMBR-T-base training dataset and sharing only around 58% of medical codes, the pretrained model achieved superior discriminative performance (AUROC ranging from 0.782 for ED visits to 0.923 for readmissions). Notably, for rare events such as unplanned admissions (0.4% prevalence), CLMBR-T-base substantially improved prediction accuracy, achieving nearly triple the AUPRC compared to the count-based feature strategy (0.037 vs. 0.011).

The robustness of CLMBR-T-base in our study suggests that pretrained transformer models capture generalizable longitudinal clinical patterns beyond institution-specific coding idiosyncrasies. Our findings illustrate clear scalability benefits, as shown by greater improvements in predictive performance with increased training sample size compared to count-based models, especially for infrequent outcomes. For example, predicting unplanned admissions showed a 16.8 percentage point improvement in AUROC from 5,000 to 100,000 patients using CLMBR-T-base, whereas the improvement plateaued early for count-based features. This result aligns with previous findings that showed the superiority of pretrained models over count-based methods in few-shot learning scenarios, albeit using smaller training data and different outcomes within the same clinical setting(Wornow et al., 2023a).

We compared CLMBR-T-base against two established baseline methods, each chosen for distinct methodological reasons. First, we evaluated against handcrafted features including comorbidity, prior drug usage, and surrogate metrics of healthcare utilization. These features represent clinically interpretable variables with established relevance to the prediction task, allowing us to assess whether our embeddings capture meaningful clinical signals beyond what domain experts would naturally select (Askar et al., 2024). Second, we compared against cumulative count features that aggregate all codes in patient histories, following not only the original CLMBR-T paper methodology, but also other works that have established this as a standard benchmark for evaluating medical embeddings (Wornow et al., 2023a; Rajkomar et al., 2018). Our results show that richer embedding representations, even when trained in a different healthcare setting with distinct care pathways and coding practices, outperform both comparison methods. The hand-crafted clinically interpretable features showed the lowest performance, while the cumulative count features achieved intermediate results, with pretrained embeddings proving most effective for predicting hospital visits. This performance hierarchy aligns with established literature showing that machine learning models using larger, less interpretable feature sets typically outperform those relying on smaller sets of interpretable features, while extending this finding to cross-institutional transfer scenarios (Rajkomar et al., 2018). Although CLMBR-T-base showed superior discriminative performance, it did not provide reliable calibration directly from initial implementation. The model consistently showed higher expected calibration errors than comparison approaches, indicating that post-hoc recalibration methods, such as temperature scaling or isotonic regression, should be applied before clinical deployment. Accurate calibration is critical, as it ensures that predicted risks closely align with observed outcomes, directly impacting clinical decision-making and resource allocation Hicks et al. (2022).

Our findings offer practical guidance for healthcare institutions considering pretrained model adoption. CLMBR-T-base’s predictive advantage increased with patients’ longitudinal record depth, as the transformer’s superiority over simpler baselines expanded substantially with growing historical code counts (Figure 4). This advantage was most pronounced for ED visits and unplanned hospitalizations, which occurred less frequently and are less clinically predictable than hospital admissions and readmissions in our dataset. For patients with sparse histories (fewer than 30 codes), interpretable tree-based models maintained competitive accuracy and may be preferable in resource-constrained environments. These results indicate that institutions seeking to predict rare, costly events should fine-tune pretrained transformers, even with minimal local data, while simpler approaches suffice for common outcomes. Furthermore, strong performance was achieved with partial rather than complete alignment between local coding systems and the model’s vocabulary, supporting recent evidence that CLMBR-T models transfer effectively across institutions using less than 1% local training data and incomplete code mapping (Guo et al., 2024).

While the strengths of this study include a rigorous comparative design using a standardized data model (OMOP) and direct evaluation of transfer learning in a very different clinical context, several limitations must be considered. First, our study utilized data from a single institution, potentially limiting generalizability. Replication across diverse settings is essential to validate our findings broadly. Second, our operational definitions of hospital visits are subject to bias. Unplanned admissions relied on antecedent ED visits as a proxy, potentially underestimating directly admitted unplanned hospitalizations. Meanwhile, many hospital admissions and readmissions could have been entirely elective and therefore too easy to predict using available data, artificially inflating predictive performance. Future work should address these definitional ambiguities through better integration of clinical workflows and admission documentation. Third, we only analyzed structured EHR data; integrating other data modalities such as clinical notes and imaging data would certainly enrich patient information and could interfere with reported predictive accuracy. Fourth, the visit-level prediction design allows patients to contribute a different number of predictions during model training, which can introduce bias toward high users. While we acknowledge this issue, we believe the approach is appropriate given the intended use of the models. These models are meant to dynamically update risk predictions at each visit in order to provide actionable clinical insights. To minimize this bias, we split the data by patient, limited the number of visits each patient could contribute to model training, and reported cluster bootstrap confidence intervals, which are known to produce less optimistic estimates. Finally, the inherent “black-box” nature of transformer-based models poses interpretability challenges critical for clinical adoption. In this study the tension between interpretability and performance was clear by our comparison of few hand-crafted features to transformer vector representations (Bouwmeester et al., 2013). Future research should focus into explainability methods of such representations, such as sparse attention mechanisms or concept-level feature attribution.

## 5. Conclusion

We show that foundational models pretrained on extensive EHR data can effectively enable transfer knowledge to institutions with smaller and sparser datasets, delivering superior predictive performance for clinically important outcomes such as ED visits and hospital admissions. However, the performance and practical utility of pretrained models vary significantly depending on outcome prevalence, patient record richness, and calibration needs. Institutions aiming to predict ED visits and unplanned visits will benefit from adopting pretrained transformer models and investing in local fine-tuning and recalibration. Future research should address interpretability and clinical integration challenges, incorporate additional data modalities, and validate these findings across diverse healthcare environments to maximize the clinical impact of advanced predictive modeling.

## Appendix A. Supplementary Figures

**Figure A.5:**
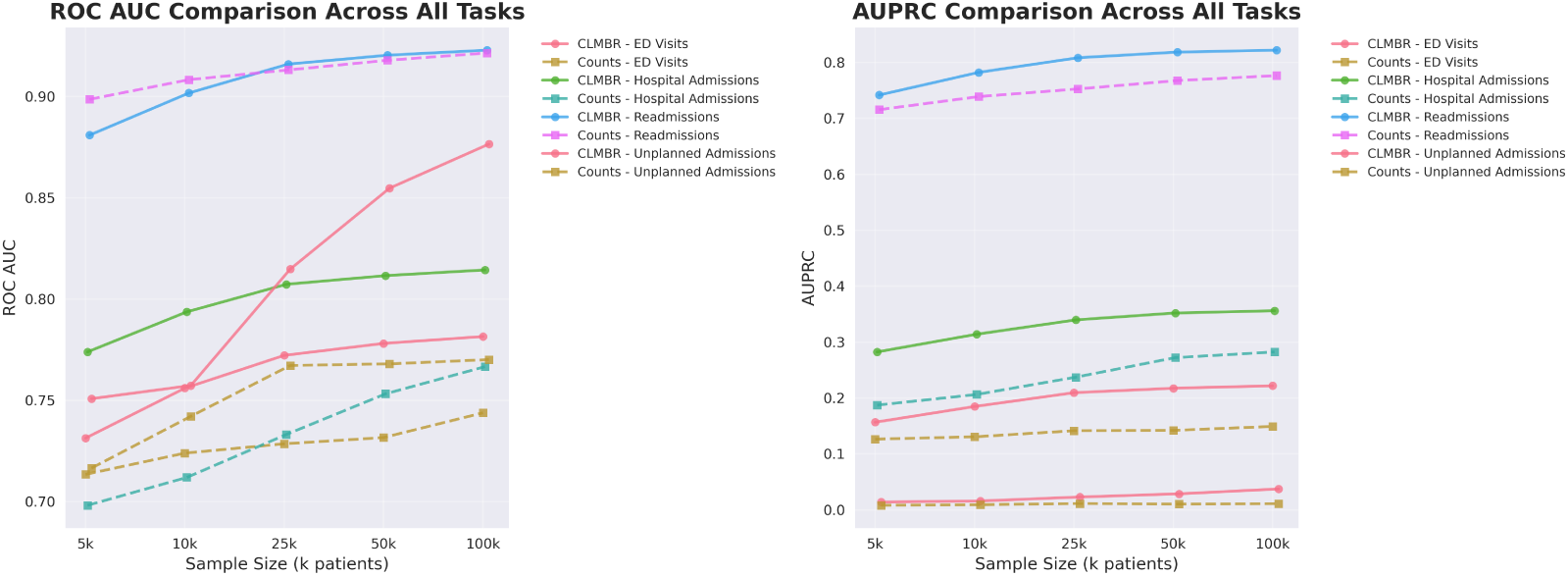
AUROC and AUPRC across CLMBR-T and Counts-based models on the four evaluated outcomes, over different sizes of traning data.

**Figure A.6:**
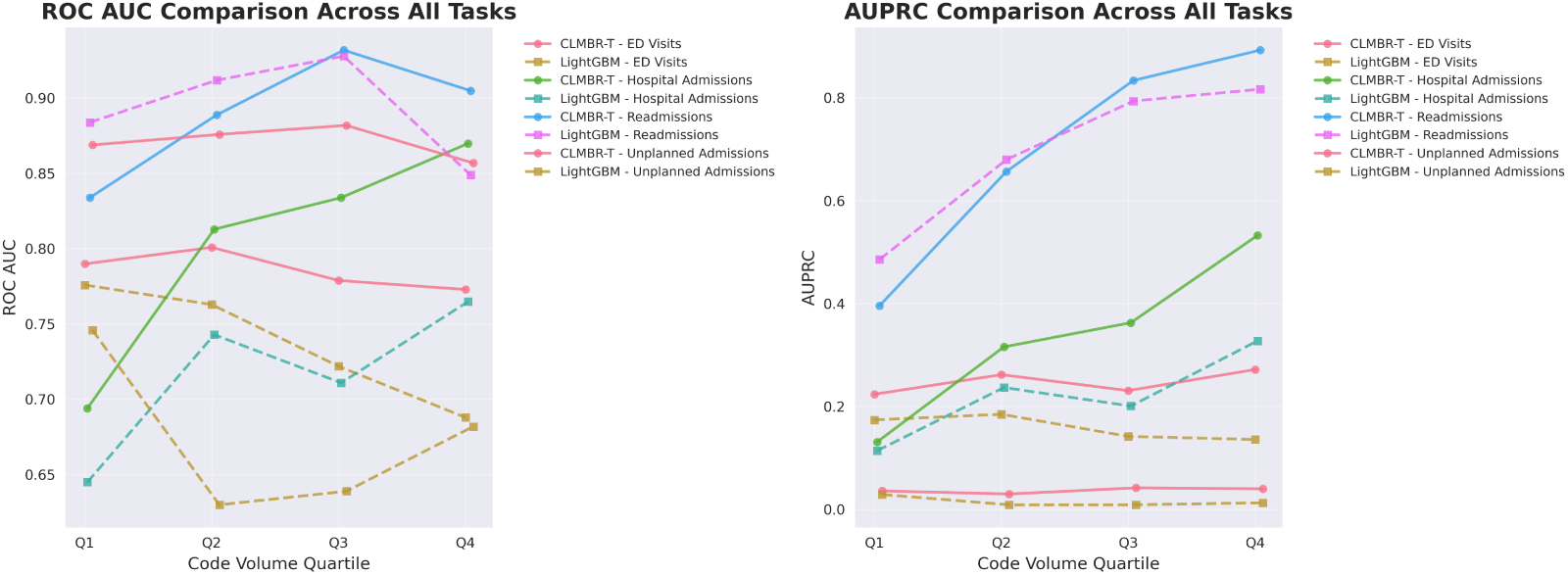
AUROC and AUPRC across CLMBR-T and Counts-based models on the four evaluated outcomes, over different quartiles of number of codes in test data.

**Figure A.7:**
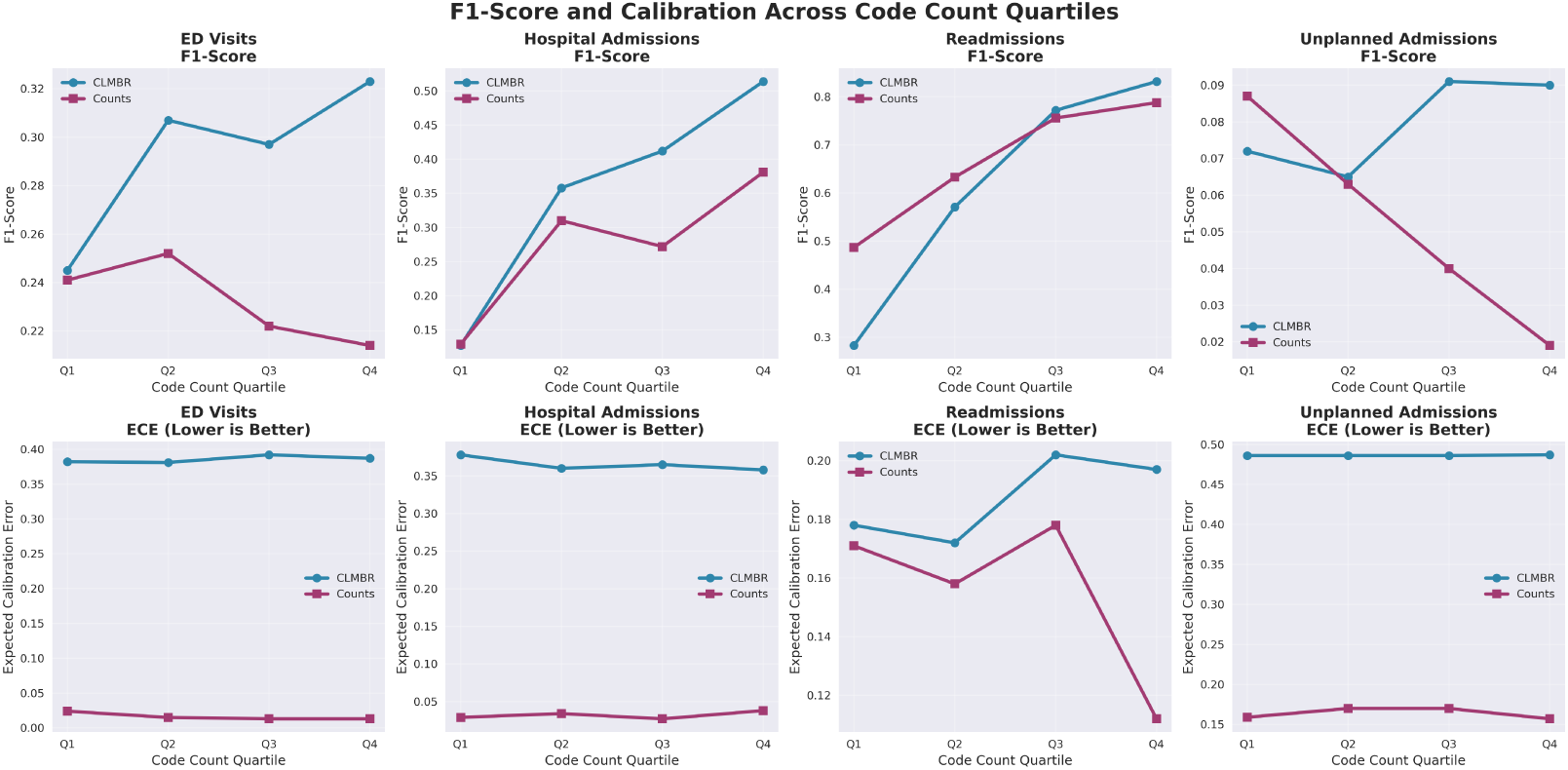
F1-score on each outcome best threshold and Estimated Calibration error (ECE) across CLMBR-T and Counts-based models on the four evaluated outcomes, over different quartiles of number of codes in test data.

**Figure A.8:**
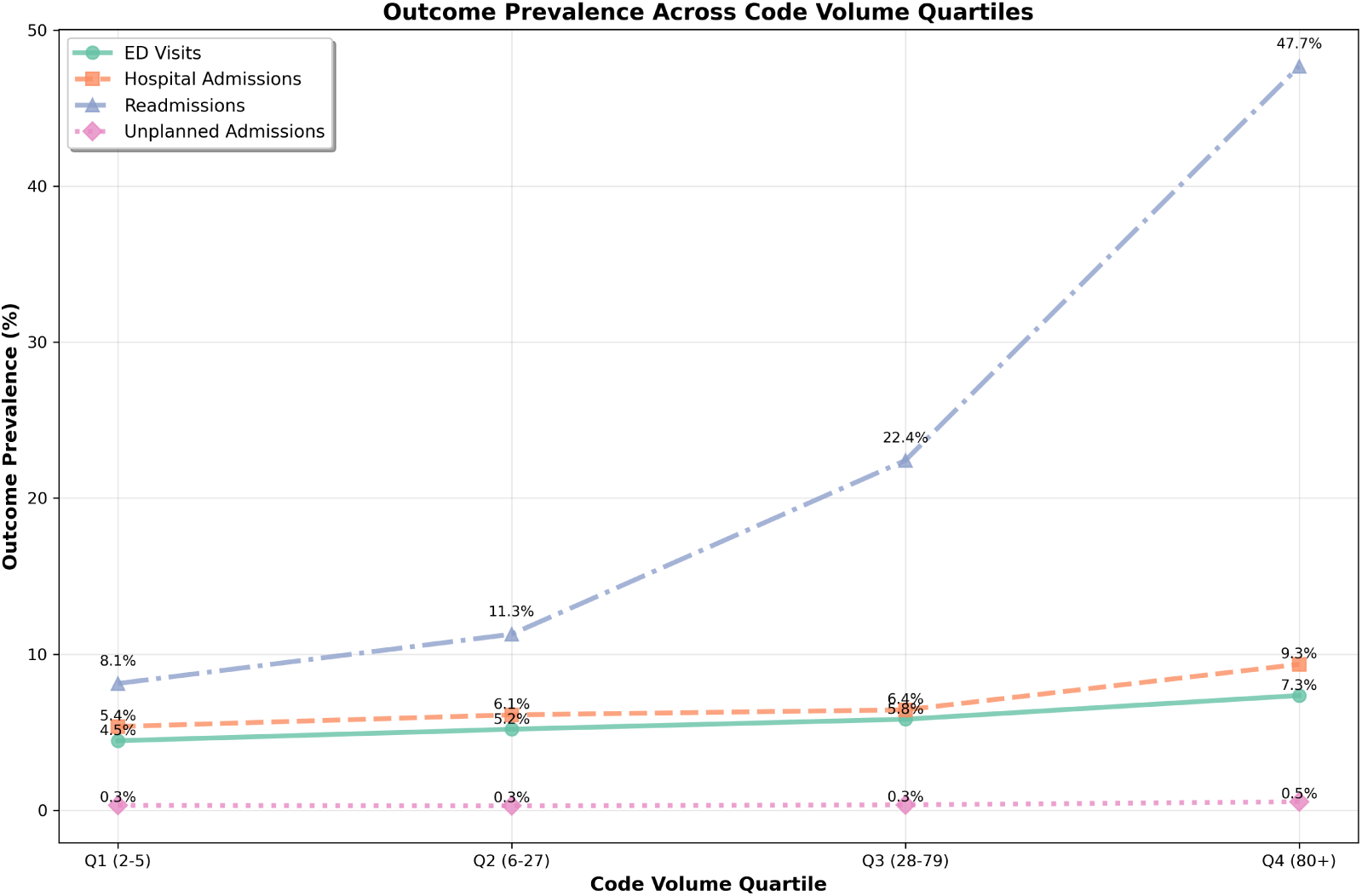
Outcome prevalence over different quartiles of number of codes in test data.

**Figure A.9:**
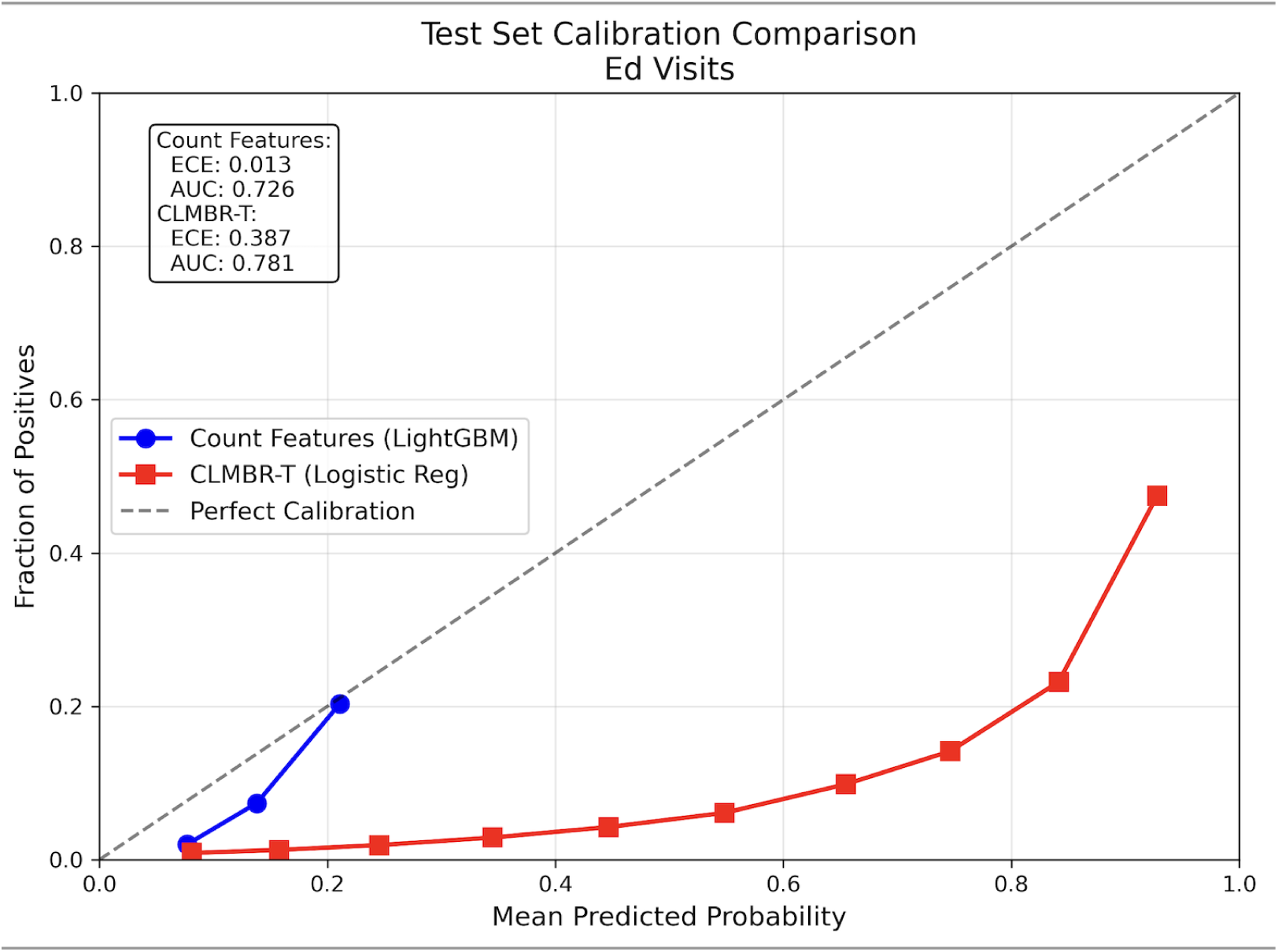
Calibration plot of ED visit prediction of Counts-based LightGBM and CLMBR-T.

**Figure A.10:**
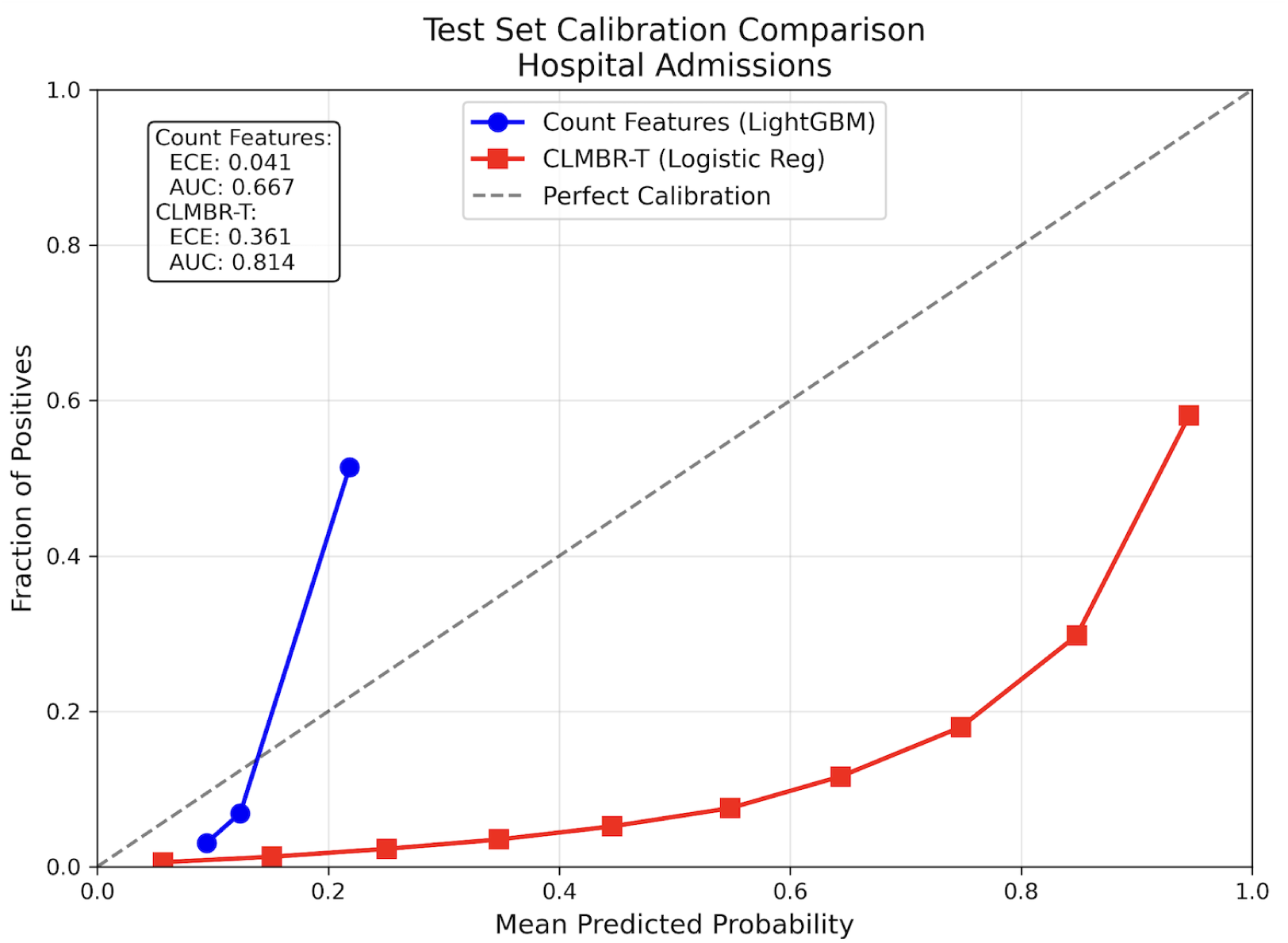
Calibration plot of Hospital Admission prediction of Counts-based LightGBM and CLMBR-T.

**Figure A.11:**
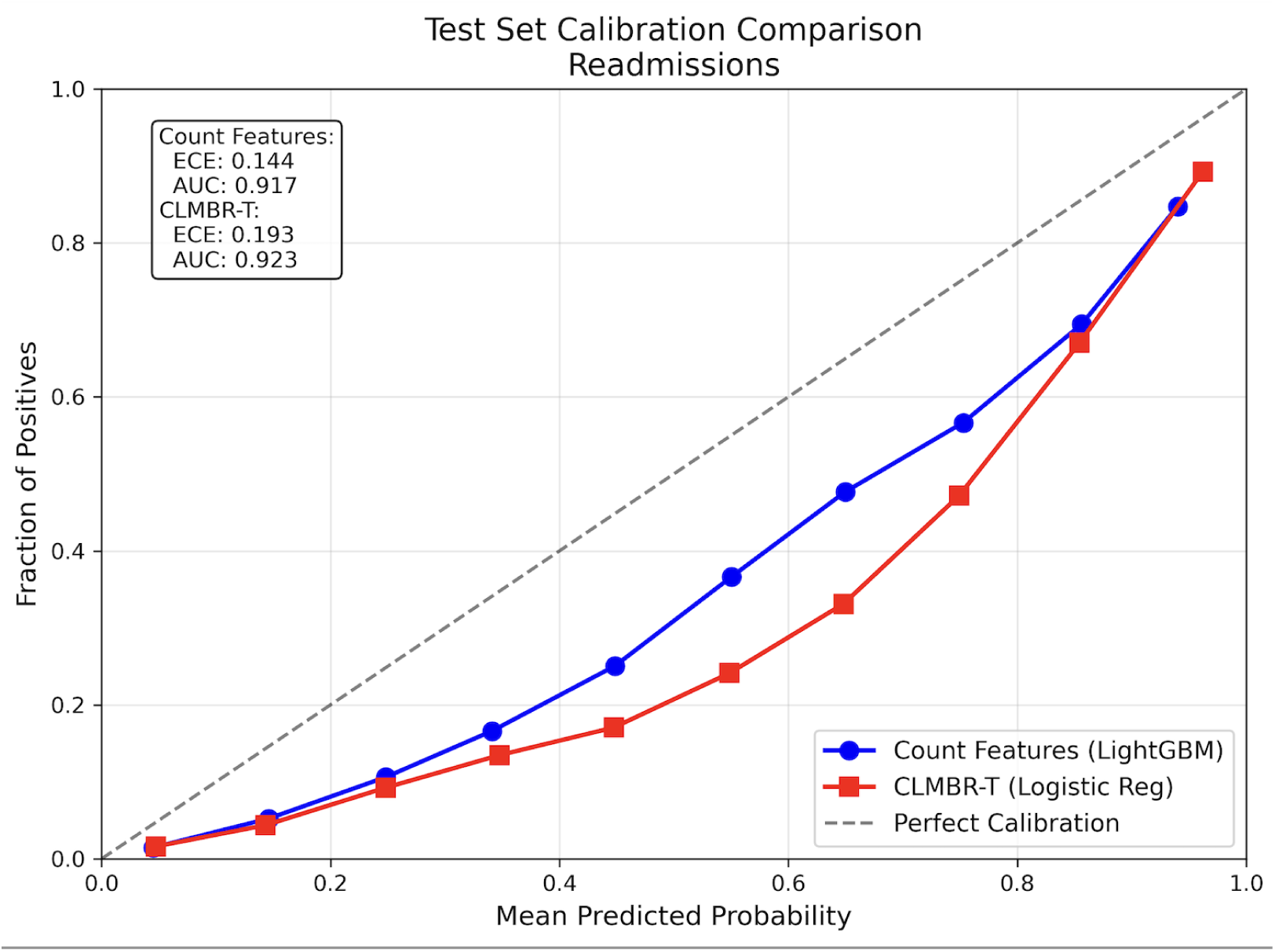
Calibration plot of Hospital Readmission prediction of Counts-based Light-GBM and CLMBR-T.

**Figure A.12:**
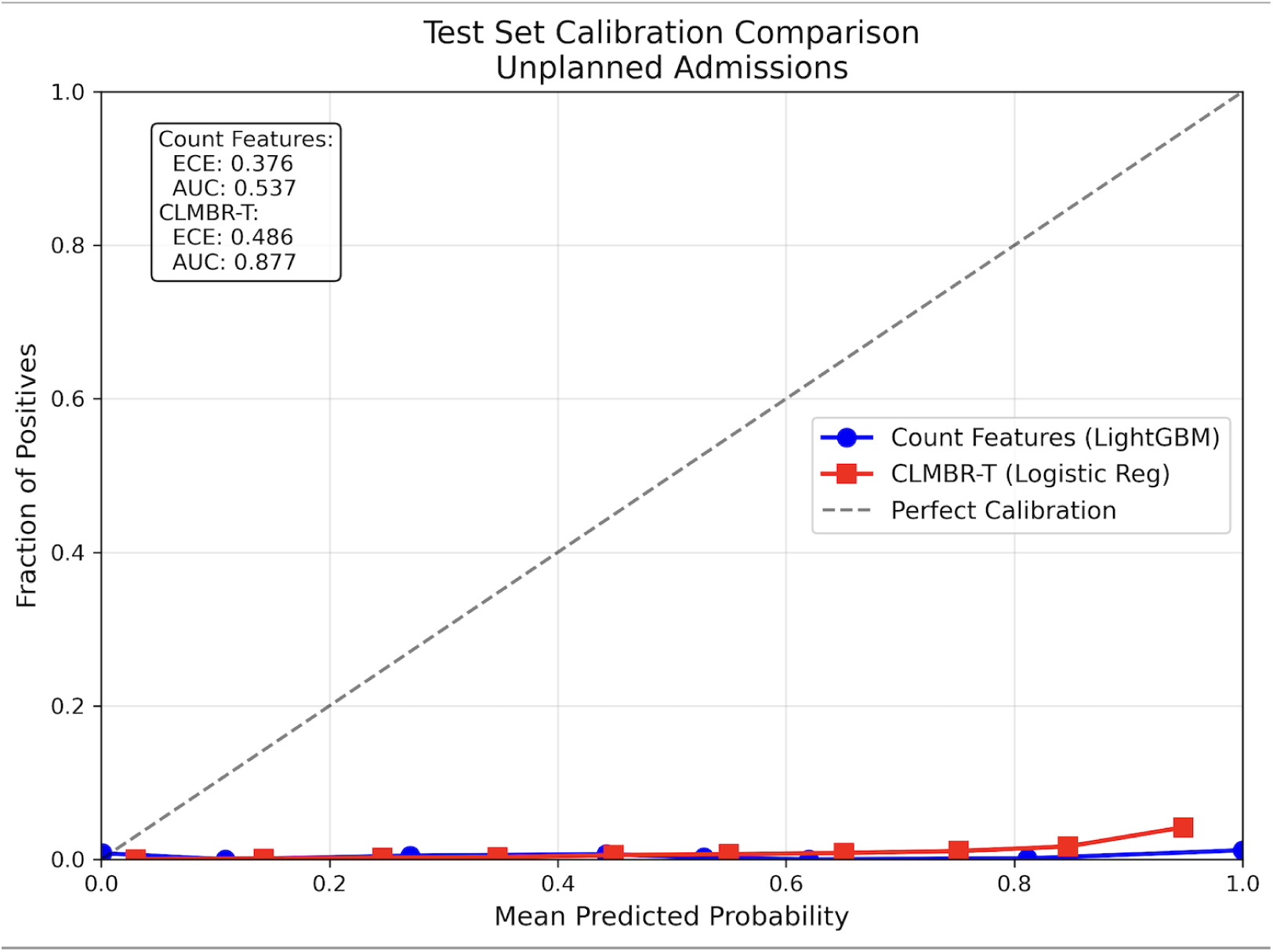
Calibration plot of Unplanned Admission prediction of Counts-based Light-GBM and CLMBR-T

## Appendix B. Supplementary Tables

**Table B.6:**
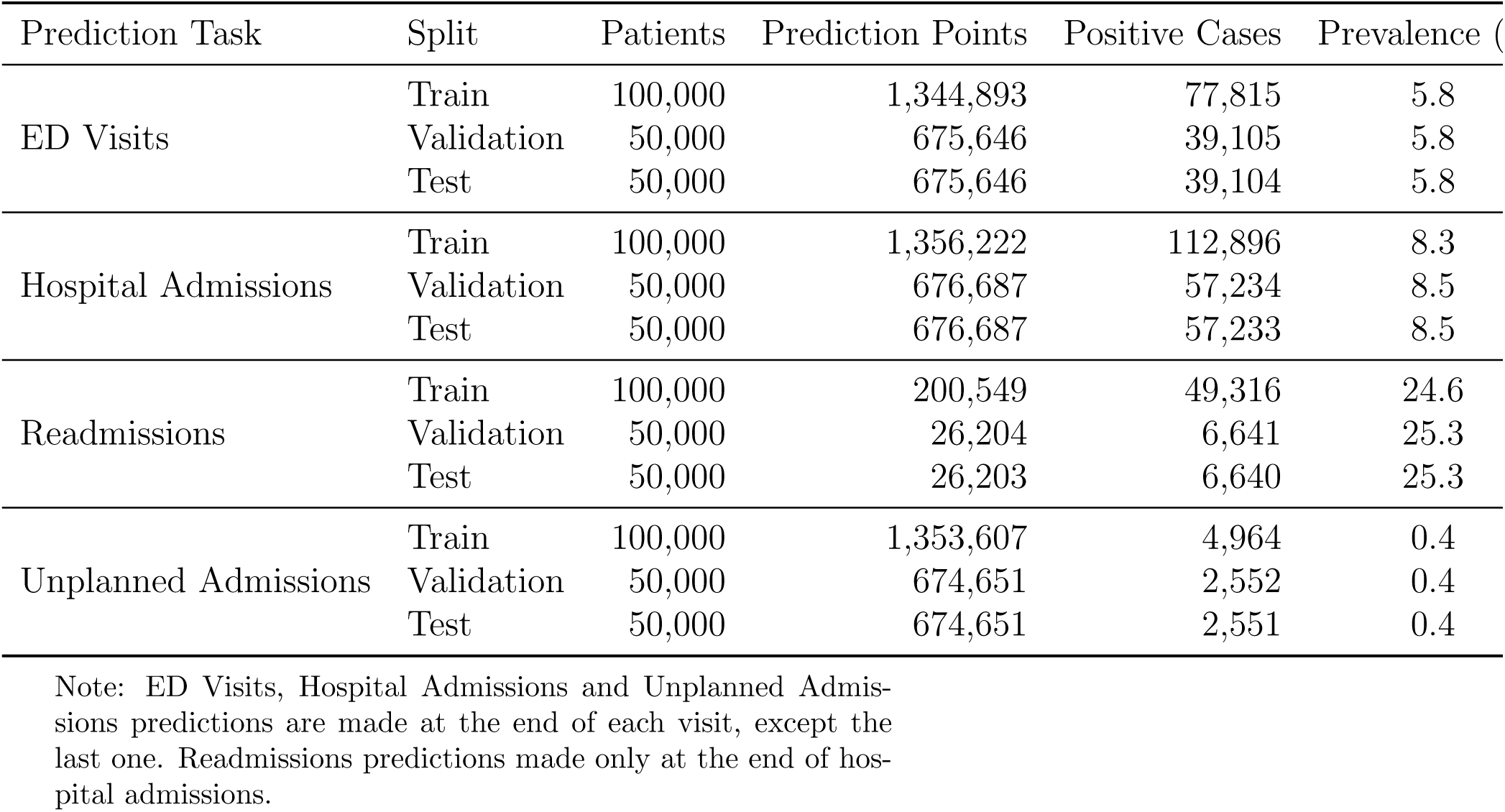
Study Population and Outcome Prevalence by Prediction Task Across Train/-Validation/Test Splits.

**Table B.7:** Extended Performance Metrics and Model Characteristics (Test Set).

| Outcome | Metric | Handcrafted | Counts | CLMBR-T |
| --- | --- | --- | --- | --- |
| ED Visit | ECE | 0.001 | 0.002 | 0.387 |
|  | MCE | 0.009 | 0.015 | 0.609 |
|  | Brier Score | 0.049 | 0.048 | 0.183 |
|  | Accuracy | 0.599 | 0.854 | 0.904 |
|  | Optimal Threshold | 0.065 | 0.078 | 0.755 |
| Hospital Admission | ECE | 0.001 | 0.356 | 0.361 |
|  | MCE | 0.008 | 0.541 | 0.568 |
|  | Brier Score | 0.066 | 0.159 | 0.168 |
|  | Accuracy | 0.554 | 0.903 | 0.918 |
|  | Optimal Threshold | 0.081 | 0.716 | 0.765 |
| Readmission | ECE | 0.048 | 0.171 | 0.193 |
|  | MCE | 0.145 | 0.264 | 0.317 |
|  | Brier Score | 0.155 | 0.103 | 0.103 |
|  | Accuracy | 0.718 | 0.878 | 0.897 |
|  | Optimal Threshold | 0.340 | 0.610 | 0.741 |
| Unplanned Admission | ECE | 0.075 | 0.125 | 0.486 |
|  | MCE | 0.326 | 0.459 | 0.906 |
|  | Brier Score | 0.013 | 0.021 | 0.116 |
|  | Accuracy | 0.523 | 0.907 | 0.988 |
|  | Optimal Threshold | 0.337 | 0.369 | 0.968 |

**Notes:** ECE = Expected Calibration Error; MCE = Maximum Calibration Error; All metrics computed on test set using optimal thresholds from validation set.

**Table B.8:**
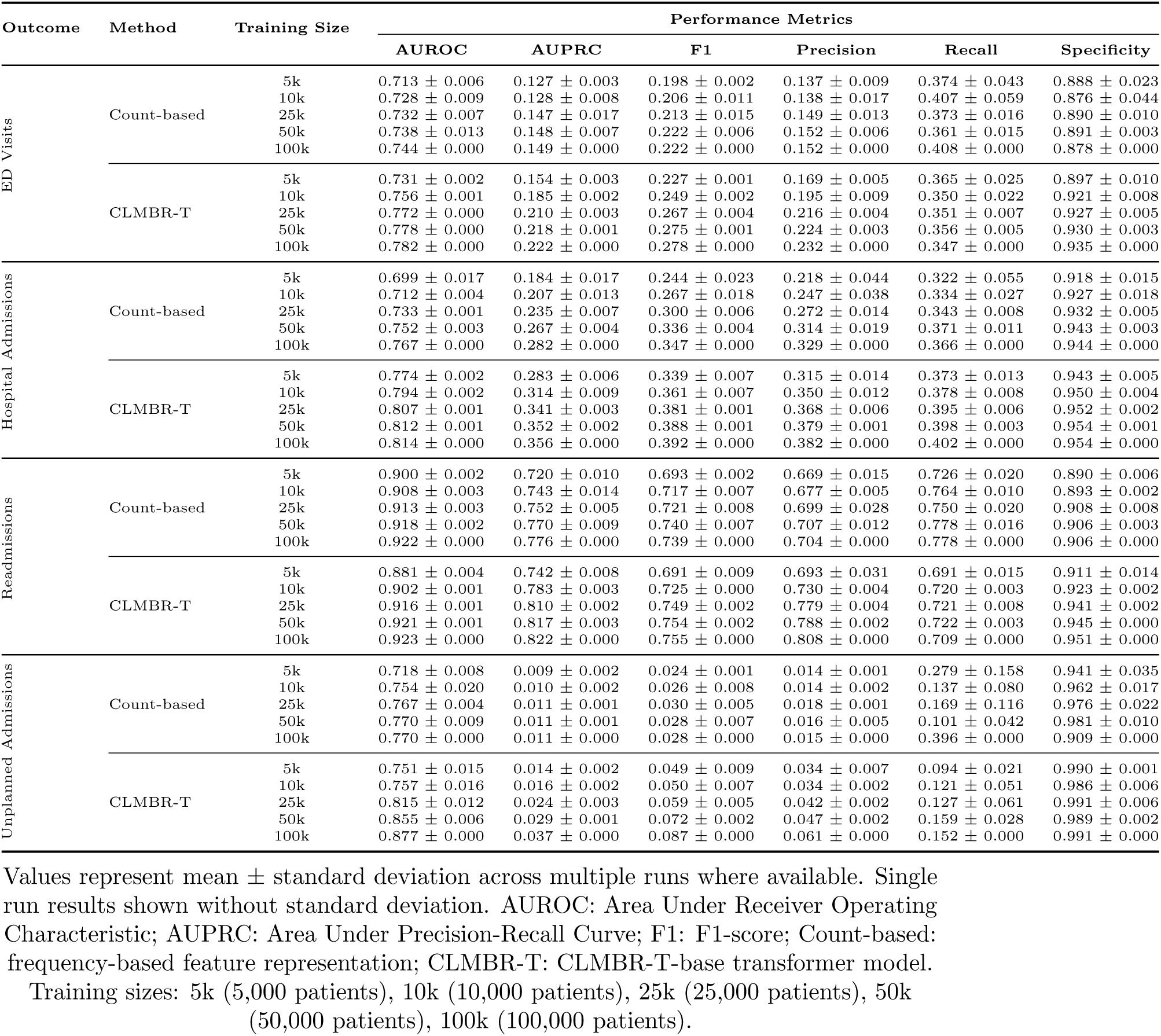
Comprehensive Performance Metrics Across Training Sample Sizes for All Prediction Tasks.

**Table B.9:** Model Calibration and Additional Performance Metrics Across Training Sample Sizes.

| Outcome | Method | Training Size | Calibration and Additional Metrics |  |  |  |  |
| --- | --- | --- | --- | --- | --- | --- | --- |
|  |  |  | ECE | MCE | Brier Score | NPV | Accuracy |
| ED Visits | Count-based | 5k | 0.010 ± 0.001 | 0.075 ± 0.006 | 0.048 ± 0.000 | 0.961 ± 0.002 | 0.849 ± 0.026 |
| | | 10k | 0.002 ± 0.000 | 0.017 ± 0.000 | 0.048 ± 0.000 | 0.964 ± 0.005 | 0.841 ± 0.034 |
| | | 25k | 0.002 ± 0.000 | 0.016 ± 0.000 | 0.048 ± 0.000 | 0.963 ± 0.001 | 0.857 ± 0.011 |
| | | 50k | 0.002 ± 0.000 | 0.016 ± 0.000 | 0.048 ± 0.000 | 0.963 ± 0.001 | 0.862 ± 0.006 |
| | | 100k | 0.002 ± 0.000 | 0.015 ± 0.000 | 0.048 ± 0.000 | 0.965 ± 0.000 | 0.854 ± 0.000 |
| | CLMBR-T | 5k | 0.411 ± 0.001 | 0.688 ± 0.003 | 0.176 ± 0.004 | 0.961 ± 0.001 | 0.865 ± 0.010 |
| | | 10k | 0.403 ± 0.000 | 0.658 ± 0.003 | 0.179 ± 0.003 | 0.961 ± 0.001 | 0.890 ± 0.007 |
| | | 25k | 0.393 ± 0.001 | 0.626 ± 0.004 | 0.182 ± 0.001 | 0.962 ± 0.001 | 0.894 ± 0.003 |
| | | 50k | 0.389 ± 0.001 | 0.616 ± 0.003 | 0.182 ± 0.000 | 0.963 ± 0.001 | 0.898 ± 0.002 |
| | | 100k | 0.387 ± 0.000 | 0.610 ± 0.000 | 0.183 ± 0.000 | 0.962 ± 0.000 | 0.904 ± 0.000 |
| Hospital Admissions | Count-based | 5k | 0.009 ± 0.001 | 0.061 ± 0.017 | 0.064 ± 0.000 | 0.948 ± 0.006 | 0.878 ± 0.025 |
| | | 10k | 0.011 ± 0.001 | 0.075 ± 0.011 | 0.064 ± 0.000 | 0.950 ± 0.005 | 0.886 ± 0.013 |
| | | 25k | 0.014 ± 0.000 | 0.093 ± 0.004 | 0.064 ± 0.000 | 0.950 ± 0.002 | 0.891 ± 0.004 |
| | | 50k | 0.354 ± 0.002 | 0.533 ± 0.008 | 0.156 ± 0.001 | 0.951 ± 0.001 | 0.901 ± 0.002 |
| | | 100k | 0.356 ± 0.000 | 0.541 ± 0.000 | 0.159 ± 0.000 | 0.952 ± 0.000 | 0.903 ± 0.000 |
| | CLMBR-T | 5k | 0.381 ± 0.002 | 0.635 ± 0.009 | 0.159 ± 0.002 | 0.955 ± 0.001 | 0.906 ± 0.004 |
| | | 10k | 0.374 ± 0.004 | 0.599 ± 0.011 | 0.165 ± 0.005 | 0.956 ± 0.001 | 0.913 ± 0.003 |
| | | 25k | 0.366 ± 0.001 | 0.578 ± 0.002 | 0.167 ± 0.002 | 0.956 ± 0.001 | 0.916 ± 0.001 |
| | | 50k | 0.362 ± 0.000 | 0.574 ± 0.003 | 0.167 ± 0.001 | 0.957 ± 0.000 | 0.918 ± 0.000 |
| | | 100k | 0.361 ± 0.000 | 0.568 ± 0.000 | 0.168 ± 0.000 | 0.958 ± 0.000 | 0.918 ± 0.000 |
| Readmissions | Count-based | 5k | 0.036 ± 0.005 | 0.073 ± 0.017 | 0.100 ± 0.001 | 0.915 ± 0.008 | 0.858 ± 0.003 |
| | | 10k | 0.061 ± 0.013 | 0.095 ± 0.014 | 0.097 ± 0.001 | 0.922 ± 0.007 | 0.866 ± 0.001 |
| | | 25k | 0.135 ± 0.001 | 0.189 ± 0.001 | 0.103 ± 0.000 | 0.925 ± 0.008 | 0.875 ± 0.001 |
| | | 50k | 0.153 ± 0.002 | 0.225 ± 0.010 | 0.103 ± 0.000 | 0.930 ± 0.003 | 0.876 ± 0.001 |
| | | 100k | 0.171 ± 0.000 | 0.264 ± 0.000 | 0.104 ± 0.000 | 0.935 ± 0.000 | 0.878 ± 0.000 |
| | CLMBR-T | 5k | 0.194 ± 0.005 | 0.334 ± 0.011 | 0.118 ± 0.003 | 0.911 ± 0.008 | 0.863 ± 0.008 |
| | | 10k | 0.189 ± 0.002 | 0.306 ± 0.016 | 0.109 ± 0.002 | 0.921 ± 0.007 | 0.878 ± 0.001 |
| | | 25k | 0.191 ± 0.007 | 0.322 ± 0.016 | 0.104 ± 0.002 | 0.923 ± 0.005 | 0.891 ± 0.002 |
| | | 50k | 0.192 ± 0.002 | 0.328 ± 0.007 | 0.103 ± 0.000 | 0.924 ± 0.004 | 0.894 ± 0.001 |
| | | 100k | 0.193 ± 0.000 | 0.317 ± 0.000 | 0.103 ± 0.000 | 0.919 ± 0.000 | 0.897 ± 0.000 |
| Unplanned Admissions | Count-based | 5k | 0.195 ± 0.150 | 0.611 ± 0.190 | 0.024 ± 0.006 | 0.997 ± 0.000 | 0.929 ± 0.027 |
| | | 10k | 0.215 ± 0.062 | 0.636 ± 0.078 | 0.025 ± 0.001 | 0.997 ± 0.000 | 0.951 ± 0.017 |
| | | 25k | 0.135 ± 0.024 | 0.469 ± 0.067 | 0.022 ± 0.005 | 0.997 ± 0.000 | 0.967 ± 0.021 |
| | | 50k | 0.149 ± 0.025 | 0.523 ± 0.067 | 0.021 ± 0.003 | 0.997 ± 0.000 | 0.971 ± 0.016 |
| | | 100k | 0.125 ± 0.000 | 0.459 ± 0.000 | 0.021 ± 0.000 | 0.998 ± 0.000 | 0.907 ± 0.000 |
| | CLMBR-T | 5k | 0.481 ± 0.003 | 0.945 ± 0.006 | 0.018 ± 0.002 | 0.997 ± 0.000 | 0.984 ± 0.003 |
| | | 10k | 0.483 ± 0.001 | 0.943 ± 0.002 | 0.036 ± 0.002 | 0.997 ± 0.000 | 0.976 ± 0.061 |
| | | 25k | 0.486 ± 0.000 | 0.933 ± 0.002 | 0.083 ± 0.000 | 0.997 ± 0.000 | 0.985 ± 0.006 |
| | | 50k | 0.487 ± 0.000 | 0.919 ± 0.002 | 0.108 ± 0.003 | 0.997 ± 0.000 | 0.984 ± 0.002 |
| | | 100k | 0.486 ± 0.000 | 0.906 ± 0.000 | 0.116 ± 0.000 | 0.997 ± 0.000 | 0.988 ± 0.000 |

Values represent mean ± standard deviation across multiple runs where available. ECE = Expected Calibration Error; MCE = Maximum Calibration Error; NPV = Negative Predictive Value. Lower ECE, MCE, and Brier scores indicate better calibration. Count-based = frequency-based feature representation; CLMBR-T = CLMBR-T-base transformer model.

**Table B.10:** Detailed Vocabulary Distribution Analysis.

| Medical Vocabulary | CLMBR-T | Our Dataset |
| --- | --- | --- |
| *Shared Code Distribution (4,489 codes in both datasets)* |  |  |
| SNOMED (clinical terms) | 4,062 (90.5%) |  |
| LOINC (lab tests) | 311 (6.9%) |  |
| RxNorm (medications) | 110 (2.5%) |  |
| Visit (encounter types) | 3 (0.1%) |  |
| Other vocabularies | 3 (0.1%) |  |
| *Dataset-Specific Code Distribution (codes unique to each dataset)* |  |  |
| SNOMED (clinical terms) | 8,095 (37.4%) | 1,938 (60.1%) |
| LOINC (lab tests) | 4,083 (18.9%) | 380 (11.8%) |
| RxNorm (medications) | 4,568 (21.1%) | 763 (23.7%) |
| CPT4 (procedures) | 3,647 (16.8%) | – |
| ATC (drug classification) | – | 131 (4.1%) |
| CARE_SITE (locations) | 396 (1.8%) | – |
| RxNorm Extension | 254 (1.2%) | 11 (0.3%) |
| ICD10PCS (procedures) | 233 (1.1%) | – |
| ICD9Proc (procedures) | 196 (0.9%) | – |
| HCPCS (procedures/supplies) | 54 (0.2%) | – |
| ICDO3 (oncology) | 52 (0.2%) | – |
| CVX (vaccines) | 41 (0.2%) | – |
| Other vocabularies | 128 (0.6%) | 0 (0.0%) |
| **Total unique codes** | **21,656** | **3,223** |
| *Complete Code Distribution (total counts including shared + unique)* |  |  |
| SNOMED (clinical terms) | 12,157 | 6,000 |
| LOINC (lab tests) | 4,394 | 691 |
| RxNorm (medications) | 4,678 | 873 |
| CPT4 (procedures) | 3,647 | – |
| ATC (drug classification) | – | 131 |
| CARE_SITE (locations) | 396 | – |
| RxNorm Extension | 255 | 12 |
| ICD10PCS (procedures) | 233 | – |
| ICD9Proc (procedures) | 196 | – |
| HCPCS (procedures/supplies) | 54 | – |
| ICDO3 (oncology) | 52 | – |
| CVX (vaccines) | 41 | – |
| Visit (encounter types) | 6 | 3 |
| Other vocabularies | 136 | 2 |
| **Total vocabulary** | **26,145** | **7,712** |

*Note:* Shared codes appear in both datasets. Dataset-specific codes are unique to each dataset. Complete distribution includes both shared and unique codes. Percentages in the shared section reflect proportions within the 4,489 overlapping codes. Percentages in the dataset-specific section reflect proportions within each dataset’s unique codes. Medical vocabulary systems: SNOMED CT (clinical terminology), LOINC (laboratory data), RxNorm (medications), CPT4 (procedures), ATC (drug classification), HCPCS (health-care procedures), ICD (diagnostic/procedure codes), CVX (vaccines), ICDO3 (oncology). CLMBR-T data from (Wornow et al., 2023a).

## Data Availability

The data used in this study, while obtained from anonymized Electronic Health Records, are subject to strict confidentiality and privacy regulations. Data access requests may be considered on a case-by-case basis and will require approval from the relevant institutional review boards and data custodians. Researchers interested in obtaining access to the data for the purpose of validating or extending the findings presented in this paper should contact the corresponding author for further information on the data access process and the necessary legal and ethical requirements. All code developed for this study will be shared publicly.

## Footnotes

1 https://github.com/som-shahlab/femr

2 https://github.com/Medical-Event-Data-Standard/medsetl

3 https://huggingface.co/StanfordShahLab/clmbr-t-base

